# Detection and characterization of Hepatitis B virus double-stranded linear DNA-derived covalently closed circular DNA in chronic hepatitis B patients

**DOI:** 10.1101/2025.01.20.25320854

**Authors:** Hsin-Ni Liu, Elena Kim, Zhili Wang, ThiThuyTu Nguyen, Fwu-Shan Shieh, Yuanjie Liu, Marc G. Ghany, Raymond T. Chung, Richard K. Sterling, Selena Y Lin, Haitao Guo, Daryl T-Y Lau, Ying-Hsiu Su

**Author notes:** Corresponding author: Ying-Hsiu Su, The Baruch S. Blumberg Research Institute, 3805 Old Easton Rd, Doylestown, PA, 18902. Authors with equal contributions (Haitao Guo, Daryl T-Y Lau, Ying-Hsiu Su). **Conflict of interest**: SL, ZW, and YS: Shareholder of JBS Science Inc. SL, FS, and ZW are employees of JBS Science, Inc at the time of the study. All other authors declare no competing interests. **Authors contributions**: YS, DL, and HG conceived the study. HL, EK, TN, and YL performed the experiments. HL, YS, SL, FS, YL, and ZW analyzed the data. MG, RC, RS, HG, and DL provided the clinical specimens and support for clinical analysis. HL, YS, DL, and HG wrote the manuscript. All authors have reviewed the manuscript.

## Abstract

**Background and Aims:** Hepatitis B virus (HBV) generates a double-stranded linear DNA (dslDNA) byproduct during replication. This dslDNA can undergo intermolecular and intramolecular nonhomologous end-joining (NHEJ) recombination, resulting in viral integration and dslDNA derived covalently closed circular DNAs (dsl-cccDNAs), respectively. The insertions and deletions (INDELs) at the end-joining site around the direct repeat (DR) 1 motif have been used to differentiate dsl-cccDNA from the authentic cccDNA. The frequency and characteristics of dsl-cccDNA in chronic hepatitis B (CHB) patients remain unclear.

**Methods:** HBV-targeted next-generation sequencing (NGS) was used to identify 32 dsl-cccDNA positive candidates, 22 HBeAg(+) and 10 HBeAg(-), from 56 liver biopsies of antiviral treatment-naive CHB patients for dsl-cccDNA confirmation and characterization by cccDNA-PCR NGS. INDELs within the DR2-1 region (nt 1600–1840) of the cccDNA were analyzed.

**Results:** Various clonally expanded, heterogenous ∼22-nt deletions in the X-gene were detected in all 32 samples, which are likely quasi-species from the authentic cccDNA. We, therefore, defined dsl-cccDNA only by the presence of INDELs clustered at the DR1 surrounding region (nt 1800–1840). The percentage of dsl-cccDNA in total cccDNA was higher among HBeAg(+) compared to HBeAg(-) samples [11.32 (3.24–26.94)% *vs.* 7.72 (2.16–28.23)%, p=0.01]. The diversity of dsl-cccDNA species correlated with cccDNA levels (log-transformed; r=0.82, p<0.001), HBeAg(+) CHB (p<0.001), and serum HBV DNA (p<0.001).

**Conclusions:** dsl-cccDNA is more prevalent and heterogenous among the HBeAg(+) CHB subjects, which is likely due to the active viral cccDNA transcription and reverse transcription at the HBeAg(+) phase. The existence of replication-defective dsl-cccDNA may facilitate immune evasion and HBV integration, and complicate HBV pathogenesis. The potential impact of dsl-cccDNA in HBV therapeutic response deserves further assessment.

## Introduction

Human hepatitis B virus (HBV), an orthotype of hepadnavirusese, is the major cause of viral hepatitis, cirrhosis, and hepatocellular carcinoma (HCC) [1]. Upon infection of hepatocytes, the initiation of hepadnaviral DNA synthesis begins with the conversion of genomic relaxed circular DNA (rcDNA) of the infecting virus to covalently closed circular DNA (cccDNA) in the nucleus of the infected cell [2]. It is known that hepadnavirus utilizes a sophisticated host repair mechanism(s) to repair both minus and plus-strands to form most of cccDNA from rcDNA [3–5]. This rcDNA-derived cccDNA is known as authentic cccDNA that serves as the template for all HBV transcripts including the pregenomic RNA (pgRNA), the template of replication, and enables viral persistence. However, a small portion of cccDNA can be formed *via* self-ligation of the viral double-stranded linear DNA (dslDNA), a byproduct of HBV reverse transcription due to an aberrant priming of the second strand DNA synthesis at the direct repeat 1 (DR1) instead of DR2 [3, 6]. Similar to the dslDNA integration into host genome, this episomal dslDNA self-ligation process is also catalyzed by the cellular error-prone non-homologues end joining (NHEJ) pathway that often inserts or deletes various length of nucleotides (INDELs) at the joint site of the dslDNA-derived cccDNA (dsl-cccDNA) [3, 7, 8], thus, hepadnaviral dsl-cccDNA species are often characterized by extensive and heterogenous INDELs at the joint site [7–12].

Due to the INDELS, dsl-cccDNA species are mostly defective of supporting new rounds of viral rcDNA replication, some can remain functional for viral DNA synthesis through multiple generations of dslDNA and dsl-cccDNA intermediates in a process called illegitimate replication by Yang and Summers [9, 10, 12], and some may support virus antigens expression as the authentic cccDNA does. Although the existence of hepadnaviral dsl-cccDNA has been demonstrated in cell cultures and duck and woodchuck animal models [7–12], the prevalence of dsl-cccDNA has not been investigated in patients with chronic hepatitis B (CHB). Due to its implications for the HBV life cycle and its perpetuation, we sought to determine whether the dsl-cccDNA species exist in CHB patients and correlate it with other HBV parameters. Interestingly, we detected dsl-cccDNA in all 32 patients studied by next generation sequencing (NGS), and it is significantly more abundant among the patients with HBeAg(+) CHB, independent of the levels of their viremia. This study is the first report that detected and characterized the differential profiles of dsl-cccDNA in livers from HBeAg(+) and (-) CHB patients. We also discovered a high prevalent newly discovered 22-nt deletion (nt 1755–1776) in HBx gene as part of authentic cccDNA. The implication of the findings in HBV pathogenesis is discussed.

## Materials and Methods

### Study subjects

Patient tissue specimens were collected by The Hepatitis B Research Network (HBRN), a research network of 28 clinical sites throughout the U.S. and Canada, funded by the National Institutes of Health, initiated to study the natural history of CHB and to conduct clinical trials in both children and adults. The Adult Cohort study (NCT01263587) enrolled HBsAg(+) subjects ≥18 years, between 2012 and 2017, who were not currently on antiviral therapy. The HBRN study protocols were approved by the institutional review boards (Research Ethics Board in the case of the Toronto site) of each participating institution, and each participant provided written, informed consent. For the present study, 56 [24 HBeAg(+), 32 HBeAg(-)] participants who had not been on antiviral therapy and had available liver tissues within 24 weeks of their clinical and virological assessments were included. The clinical information is summarized in **Supplemental Table 1**.

### Identification and characterization of dsl-cccDNA by HBV-targeted NGS and PSAD-cccDNA PCR NGS assays

Total DNA, previously isolated from the frozen liver biopsies, as detailed in **Supplemental Table 2**, was used. Approximately 100 ng of tissue total DNA was fragmented (100–250 bp) by sonication and subjected to library DNA preparation using the xGen™ cfDNA & FFPE DNA library prep kit (Cat# 10010207, Integrated DNA Technologies, Coralville, Iowa), which contains unique molecular identifies (UMIs), according to manufacturer specifications. Library DNA was subjected to the JBS HBV-targeted NGS assay following the manufacturer’s instructions (JBS Science Inc, Doylestown, PA). The captured library DNA was pooled for NGS analysis on the Illumina NGS platform for 2×151 bp sequencing. Sequencing reads were processed and analyzed by using a commercial pipeline JBS *Advanced ChimericSeq* version (JBS Science). Sequences that passed the pre-processing steps were aligned to a combined reference of human genome GRCh38.p13 and 42 HBV genomic sequences [13] by BWA-MEM [14]. Consensus sequences were generated for each UMI using fgbio CallMolecularConsensusReads http://fulcrumgenomics.github.io/fgbio/ [15]. An additional alignment to the HBV references was applied to extract HBV reads.

As outlined in **Figure 1**, to select samples for plasmid-safe ATP-dependent (PSAD)-cccDNA PCR-NGS assay to confirm and characterize dsl-cccDNA, we first analyzed HBV-targeted NGS sequences from 56 liver biopies for INDELs in the DR2-1 region (nt 1600–1840). As INDELs generated by different self-ligation events vary and INDELs that occurred due to the sequencing errors by Illumina NGS are reported to be rare [16]; every INDEL detected is considered in the data analysis. Deletion sequences were further characterized by the number of UMI-consolidated supporting read (SR). Sequences of interest were examined with Integrative Genomics Viewer [17] to determine the likelihood of the deletion sources for being derived from dsl-cccDNA (candidates for dsl-cccDNA-positive), as the criteria described in **Figure 1**. 32 were selected for plasmid-safe ATP-dependent (PSAD)-cccDNA PCR-NGS assay for a concordance study.

**Figure 1.**
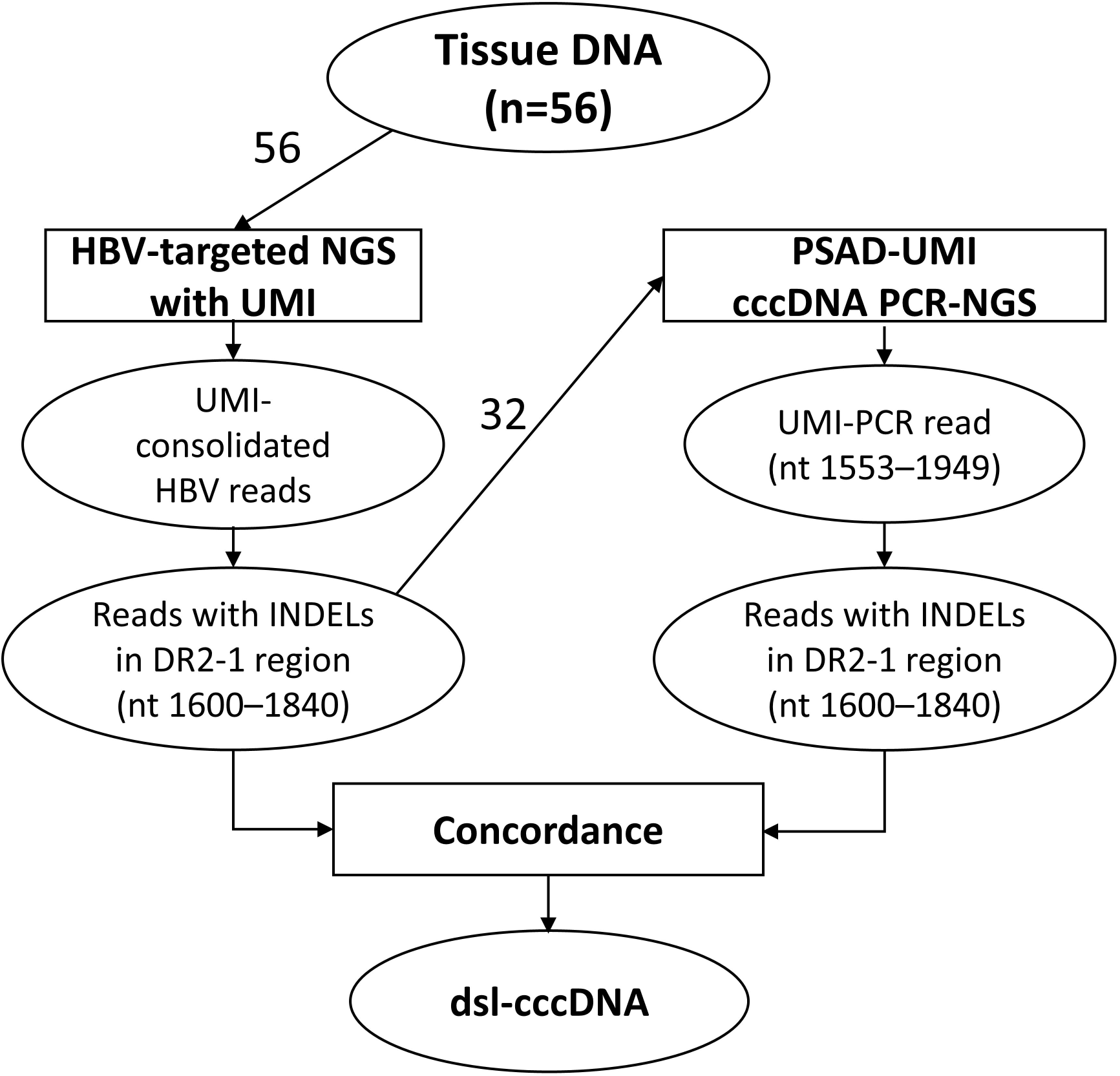
The study outline. Tissue DNA from 56 patients was subjected to HBV-targeted NGS with unique molecular index (UMI) incorporated. HBV reads were extracted by mapping reads to HBV reference genomes and the consensus sequences were generated for each UMI family. HBV reads were analyzed for INDELs in the DR2-1 region. Samples containing more than 5 different deletions and each deletion has less than 5 supporting reads were considered “dsl-cccDNA positive” by HBV-targeted NGS assay. Total of 32 samples, including 30 positives, 2 negatives for dsl-cccDNA by HBV-targeted NGS assay were selected and PCR products from PSAD-cccDNA assays were subject to PSAD-cccDNA PCR-NGS with UMI for dsl-cccDNA study. HBV reads from PSAD-UMI cccDNA PCR-NGS were analyzed for INDELs in the DR2-1 region. INDELs that were concordant between the two independent NGS assays were identified as dsl-cccDNA and were further characterized.

cccDNA PCR products generated from heat-denatured, PSAD-digested tissue DNA, as described previously [8, 18], were subjected to library preparation using xGen™ DNA Lib Prep MC UNI 96rxn (Cat# 10009820, Integrated DNA Technologies). NGS was performed by MedGenome (MedGenome Inc., Delaware, USA) on the NovaSeq at 2×250 bp.

INDELs that were found by both NGS assays were defined as concordant dsl-cccDNA INDELs. The percentage of dsl-cccDNA of total cccDNA was determined based on the concordant INDELs. INDELs in the nt 1800–1840 region detected by cccDNA PCR-NGS assay were further analyzed for INDEL diversity. The positions of all INDELs per NGS read were profiled and defined as an INDEL species. The frequencies for every INDEL species were determined for each sample, with a sum of total INDEL species equal to 100% in each sample. Shannon’s diversity index was applied to calculate INDEL species diversity based on the frequency table.

### Validation of the 22-nt HBV deletions by PCR cloning and Sanger sequencing

The sequence containing a 22-nt deletion (nt 1755–1776) identified in Pt 5 and Pt 7 was compared to previously reported HBV sequences by a similarity search in GenBank. We used genotype C (accession no. GQ377617.1) and the region of nt 1680–1798, with and without nt 1755–1776 deletion, to BLAST against NCBI nr/nt database (taxid: 10407). Only hits with 100% query coverage and deposited as complete genomes were used in the comparison. To validate the presence of the 22-nt deletion, two primer sets, one forward primer (nt 1680–1696) ATGTCAACGACCGACCT, and two reverse primers: reverse 1 CTAATACAAAGACCTTTAACCT (nt 1798–1779), and reverse 2 which is deletion-specific primer, AATTTATGCCTACAGCCTCAA (nt 1795–1753 with deleted 1776–1755), as illustrated in **Supplemental Figure 1A**, were designed based on the consensus of NGS sequences obtained from Pt 5 and Pt 7. DNA isolated from two specimens (Pt 5 and Pt 7) was used to generate PCR products using Phusion High-Fidelity DNA Polymerase (#F530S, Thermo Fisher Scientific, Waltham, MA). The dA overhangs were added using native *Taq*DNA Polymerase (#EP0282, Thermo Fisher Scientific) or LA *Taq*® DNA Polymerase (#R002M, TaKaRa, San Jose, CA). Haplotypes-containing fragments were ligated with a plasmid backbone using pGEM®-T Easy Vector System (#A137A, Promega, Madison, WI) and sequenced via pUC/M13 forward and reversed sequencing primers. Alignment was performed using MEGA 7.0.26 software [19].

### Statistical analysis

Statistical analysis and data visualization were carried out on R Studio with R version 4.2.1 (R Core Team, 2022). Wilcoxon’s Rank Sum test (R function wilcox.test) was used for comparisons of % dsl-cccDNA and INDEL diversity of dsl-cccDNA between HBeAg levels [HBeAg(+) and HBeAg(-)], and between serum HBV viral loads (<6 log IU/ml and ≥6 log IU/ml). Pearson’s correlation (R function cor.test) was used to test the relationships between % dsl-cccDNA and cccDNA level, and between INDEL diversity of dsl-cccDNA and cccDNA level. A *p*-value <0.05 was considered statistically significant.

## Results

### Detection of dsl-cccDNA by a concordance study of HBV-targeted NGS and PSAD-cccDNA-PCR NGS assays

To detect the dsl-cccDNA in liver biopsies from CHB patients, two NGS approaches, HBV-targeted by hybridization (HBV-targeted) or cccDNA PCR (PSAD-cccDNA-PCR) NGS were employed in this study, as outlined in **Figure 1**. The HBV sequences obtained from HBV-targeted NGS assay can be from all forms of HBV DNA, including cccDNA, integrated HBV DNA (iDNA), and various replicative intermediates, existing in each respective biopsy. On the other hand, the HBV sequences obtained from PSAD-cccDNA-PCR NGS are from DNA templates resisted to heat-denaturation and PSAD digestion and defined by PCR primers (nt 1553–1949) that includes the *bona fide* cccDNA and dsl-cccDNA [8, 18]. One feature of dsl-cccDNA is the existence of various INDELs at the joint site, generated by events of self-ligation, presumably within the DR2-1 region (nt 1600–1840). We first identified the tissue biopsies that most likely contain dsl-cccDNA forms by examining deletions in the DR2-1 region, as described below.

First, from HBV-targeted NGS study of 56 liver biopsies, two deletion patterns viewed by IGV were noted. The first pattern is the deletion supported by many supporting reads (SRs), as one example shown in **Figure 2A**. A 22-nt deletion, nt 1755–1776, supported by 3,356 SRs of 39,710 total HBV reads was identified in Pt 5, indicating that at least thousands double-stranded HBV DNA templates contained this 22-nt deletion in ∼100 ng of tissue biopsied DNA. The second pattern is the example shown in **Figure 2B** from Pt 4 that many deletions were supported by a single SR, suggesting each deletion was derived from one individual DNA template. We reasoned that the second pattern is more likely derived from self-ligations since each ligation is an independent event, resulting in an independent species of INDEL.

**Figure 2.**
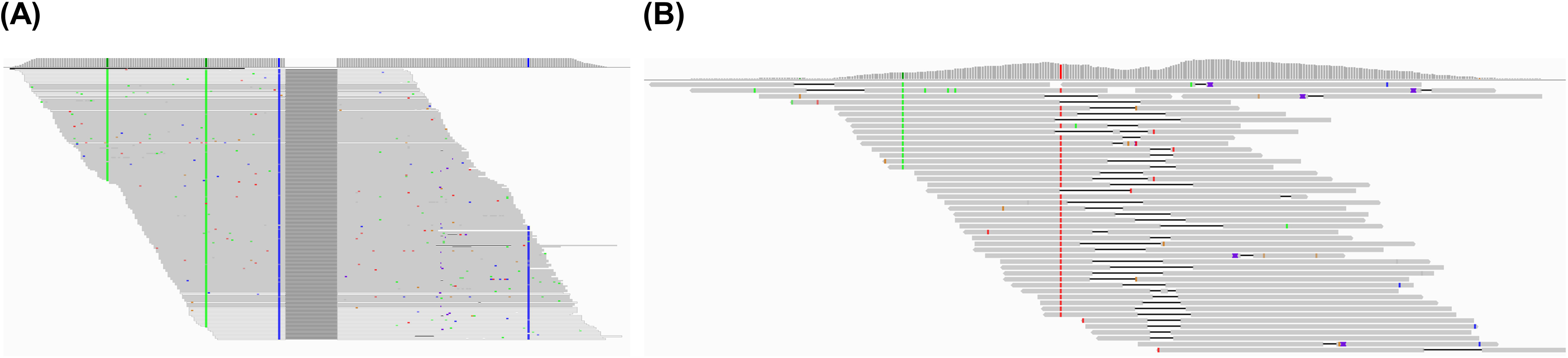
Two major deletion patterns identified in the DR2-1 (nt 1600–1840) region of the CHB tissue biopsies by HBV-targeted NGS assay. (A) An example of a 22-nt deletion at nt 1755–1776 identified in Pt 5 with 3,356 supporting reads. (B) An example of various lengths of deletions with 1 SR across nt 1684– 1957. Blue, green, and red indicate SNPs. Note: Reads in IGV snapshot was condensed and may not show all supporting reads.

Next, we selected samples that contained more than 5 different deletions in the DR2-1 region to perform deletion characterization. Each deletion should have less than 5 UMI-consolidated SRs as each dsl-cccDNA molecule should be generated by an individual self-ligation event. Two or more UMI-consolidated SRs supported for one deletion can happen when two or more self-ligation events create same deleted sequences (dsl-cccDNA) or iDNA with the deletion that expands by cell proliferation. We assumed that the same deletion created by 5 different self-ligation events should be very rare, thus <5 SRs is one of the selection criteria. Thirty-two samples, as listed in **Supplemental Table 2**, column I and J, were selected using the criteria outlined in **Figure 1**, detailed in the Material and Methods for PSAD-cccDNA-PCR NGS study, including 2 negative controls for dsl-cccDNA. Concordant INDELs detected by these two independent NGS assays were used to identify dsl-cccDNA for further characterization.

To prepare for NGS library construction, 32 PCR products from PSAD-cccDNA PCR assay was first subjected to PCR product size analysis by capillary electrophoresis. Interstingly, a significant amount of PSAD-cccDNA PCR products were shorter than the expected size of 397 bp (nt 1553–1949) for all 32 PCR products, as three examples, Pt 2, Pt 22, and Pt 41, shown in the **Supplemental Figure 2A**, indicated by arrows. A pilot NGS was performed on the PCR products of the first five patients, Pt 2, Pt 16, Pt 17, Pt 22, and Pt 41, to examine the overall composition of the PSAD-cccDNA PCR products, as summarized in the inserted table in the **Supplemental Figure 2B**. Although over one million NGS reads were obtained, over 99.9% of NGS reads do not contain both HBV cccDNA primer sequences, *i.e.* they were non-specific PCR products. Most reads were mapped to human host sequences only. We thus performed DNA size fractionation to remove PCR products less than 200 bp to enable NGS sequencing of the target HBV PCR product size range. As a result, we obtained 72.2–93.6% of NGS reads that are specific for HBV PCR reads containing both cccDNA primer sequences. The sequencing data was analyzed for INDELs in the DR2-1 (nt 1600–1840) region and then the concordance study, as summarized in **Supplemental Table 2**.

### Distribution of concordant INDELS identified by two independent NGS assays

INDELs at the joint regions, presumably the DR2-1 region, are one characteristic of dsl-cccDNA. Two analyses were performed for the INDELs concordantly identified within nt 1600–1840 from both NGS assays. We first characterized the concordant INDELs using data from PSAD-cccDNA-PCR NGS for the sizes, locations, and frequencies. The frequency of up to 30 most frequent deletions and insertions among all identified INDELs were calculated, represented by the sidebars as shown in **Supplemental Figure 3**. Two “negative” controls, as defined by less than 5 INDELs, Pt 53 and Pt 46, contained 2 and 3 qualified deletions (<5 SRs) in HBV-targeted NGS, and have 2 and 1 deletions in concordance between two NGS assays, respectively **(Figure 3A)**. Of 30 biopsies selected as positive candidates for dsl-cccDNA, three types (I–III) of INDEL distributions were identified by primary clustered locations, as individually shown in **Supplemental Figure 3** and represented by three liver biopsies in **Figure 3B**. Firstly, Pt 22 representing 9 biopsies, categorized as Type I, exhibited both insertions and deletions primarily clustered around nt 1820 near the DR1 site (nt 1824–1834). Secondly, Pt 16 representing 20 biopsies, categorized as Type II, showed deletions clustered at around nt 1760, with or without a second cluster at around nt 1820, while insertions were centered at nt 1820. Thirdly, Pt 7, the only biopsy as Type III, displayed both insertions and deletions centered at both nt 1760 and nt 1820 (**Supplemental Figure 3**). Collectively, deletions examined from all 30 biopsies were found in two clusters, but insertions were predominately concentrated around the DR1 site (nt 1820) as shown in **Figure 3C**.

**Figure 3.**
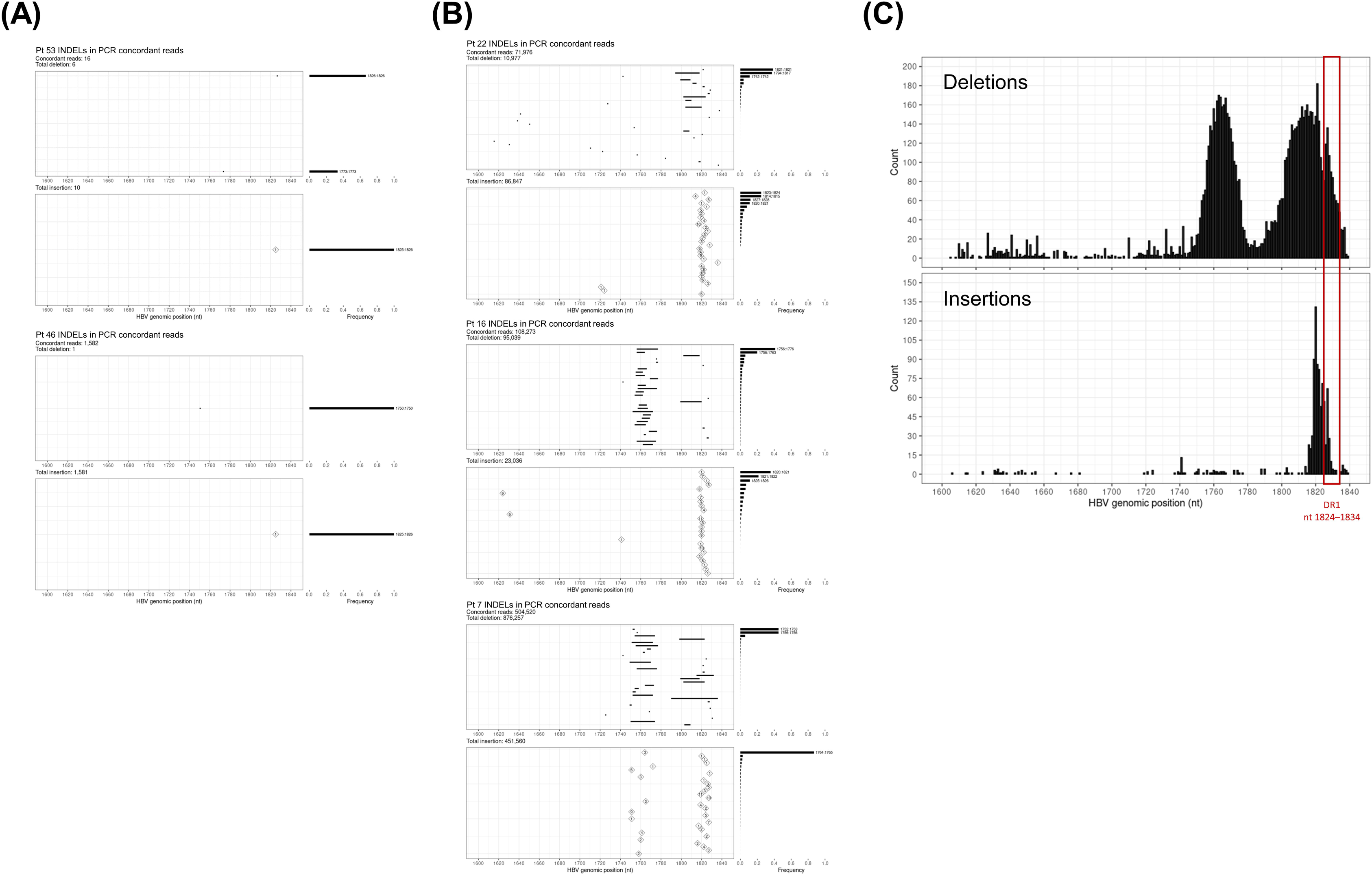
Distribution and frequency of verified (concordant) deletions and insertions (INDELs) in PSAD cccDNA PCR-NGS. Positions of each INDEL with frequencies larger than 10% are indicated. (A) INDELS detected in two negative controls. (B) Examples of three INDEL distribution patterns. (C) Distribution of overall deletion (upper) and insertion (lower) positions of INDELs detected by PSAD cccDNA PCR-NGS. Each unique INDEL was counted once in each sample, regardless of the supporting reads. The counts were the sum from 32 samples. The DR1 region (nt 1824–1834) is indicated in a red box.

### Characterization of the INDELs in the dsl-cccDNA in CHB / Discovery a 22-nt deletions in X coding region

By examining 30 most frequent INDELs distribution and their corresponding frequencies of all 30 samples, detailed in the **Supplemental Figure 3**, we noted, in some patients, there are one to few deletions composed of more than 20% of all concordant deletions detected, such as one 22-nt deletion (nt 1755–1776) from Pt 5. As illustrated in **Figure 4**, this 22-nt deletion was composed of 86% of the concordant deletions detected, and 57.4% of all detected deletions from the cccDNA-PCR NGS assay with 3,356 SRs from HBV-targeted tissue NGS in Pt 5, suggesting there were at least thousands copies of HBV cccDNA that contain this 22-nt deletion. Interestingly, the second most frequent deletion detected in Pt 5 is also in a similar region. This contradicts one of our dsl-ccsDNA selection criteria where deletions in dsl-cccDNA generated by an individual self-ligation event should be in a single type of dsl-cccDNA unless the same deletions were created from different ligation events which should be in low frequency. It was of interest to investigate the prevalence of this 22-nt deletion. Surprisingly, this 22-nt deletion was found in all 32 biopsies with various abundance detected by the PSAD-cccDNA-PCR NGS assay. Pt 7 is one example with this 22-nt deletion composed of 0.27% of all detected deletions from cccDNA PCR-NGS (**Figure 4**). We noted that for those having over 20% of INDELs were mostly around nt 1760 position, suggesting the INDELs around nt 1760 may not be from individual self-ligation events, but from amplification of cccDNA with this 22-nt deletion in the molecules *via* rcDNA recycling or *de novo* infection.

**Figure 4.**
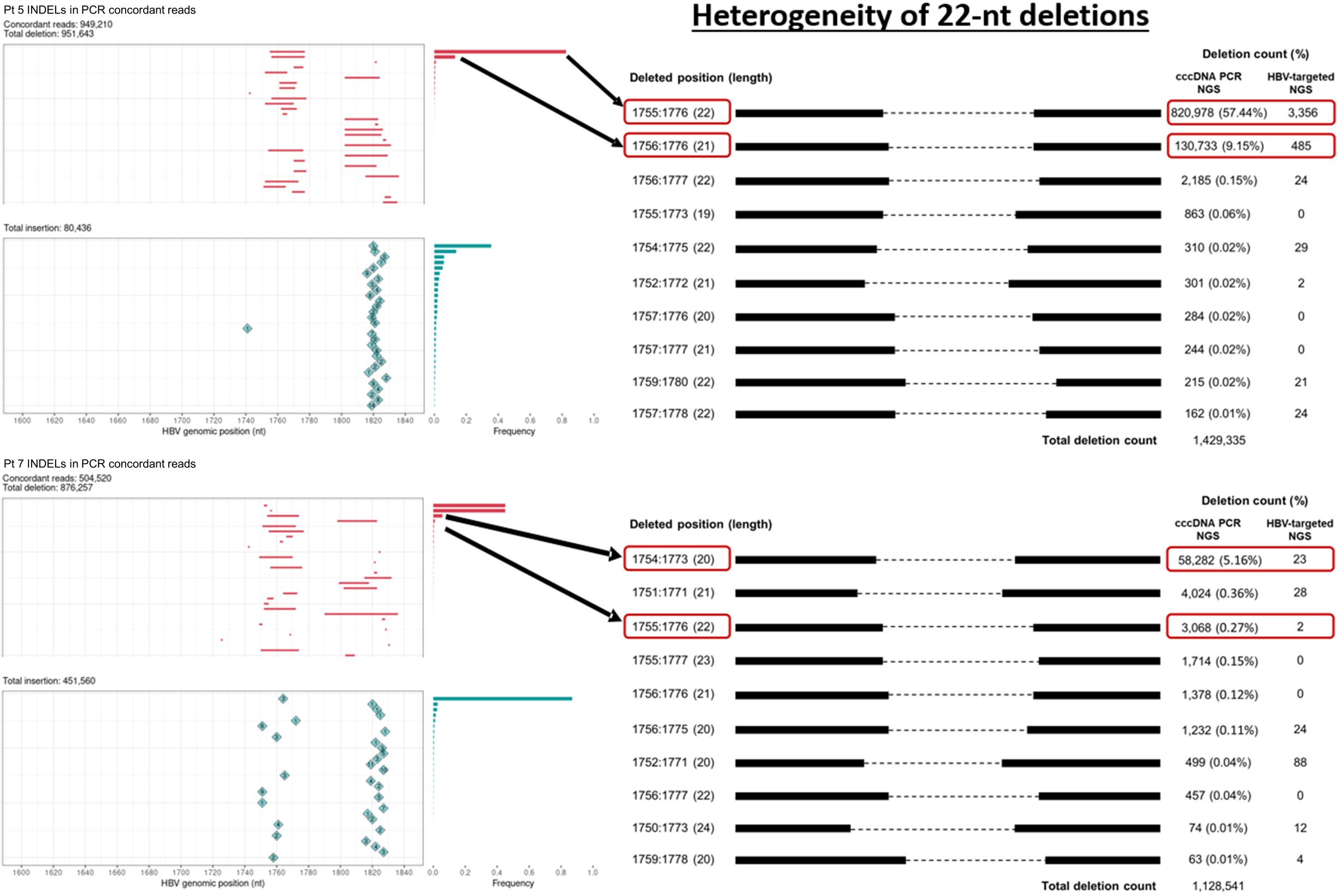
Heterogeneity of ∼22-nt deletions around nt 1755, shown by two examples, Pt 5 and Pt 7. Positions and lengths of the deletions are indicated with detection count and percentage of the total deletions detected in the region of nt 1600–1840 by both HBV-targeted and cccDNA PCR-NGS assays.

To investigate the sources of the abundant deletions around nt 1760, we first investigated if this highly abundant 22-nt deletion detected in Pt 5 has been previously reported by others. The similarity search by BLAST using deleted HBV DNA sequences from nt 1680–1798 in the GenBank was performed to report the 100 most matched sequences. As summarized in **Supplemental Table 3,** this 22-nt deletion has not been specifically reported in the GenBank or in the literature to the best of our knowledge. Interestingly, 78% of the 100 HBV DNA sequences deposited in GenBank have more than 90% identity to the sequence with deletions in this region suggesting its high prevalence. A 20-nt highly frequent heterogenous deletion (such as nt 1753–1772 or 1758–1777) that can affect X/BCP/ENII genes in CHB patients was previously reported [20] and was also identified in our study population including Pt 7 (**Figure 4)**. Next we analyzed the deletion size composition in the 32 nt region of nt 1750–1781. As shown in **Supplemental Figure 4**, 49 of 56 samples (88%) have deletions identified in this region by HBV-targeted NGS assay. 1-nt deletions are common across 47 of the 49 samples (96%), while 18–23 nt deletions were found in 14 patients (29%) in this 32 nt region.

### Validation of a newly discovered 22-nt deletion (nt 1755–1776) by PCR cloning and Sanger Sequencing

We selected two samples, Pt 5 and Pt 7, to validate the existence of this newly discovered 22-nt deletion by PCR cloning and Sanger Sequencing. This 22-nt deletion is composed of 57.4% and 0.27% of total deletions identified by cccDNA-PCR NGS in Pt 5 and Pt 7, respectively. Two sets of primers were designed for this validation study, as illustrated in **Supplemental Figure 1,** detailed in Materials and Methods. As expected, 22-nt deletions were detected in 3 of 4 (75%) clones from primer pairs for both wild-type and deleted templates, and 1 of 1 clone by deletion-specific PCR in Pt 5, and 2 of 10 (20%) clones from primers for both wild-type and deleted sequences in Pt 7, as shown in **Supplemental Figure 1B and C**. Together we conclude that this 22-nt deletion exists in all 32 of 32 CHB patients studied.

### Size distribution of INDELs occuring in nt 1800–1840

To characterize INDELs of dsl-cccDNA, we analyzed the size of each inserted and deleted sequence for all 32 cccDNA PCR NGS samples. The INDEL size distribution for each biopsied sample was plotted in the **Supplemental Figure 5** and collectively summarized in **Table 1**. Consistent with previous INDELs described from tissue cultures and animal studies [7–12], the sizes of INDELs of dsl-cccDNA were mostly less than 10 nt. 81% of insertions are shorter than 11 nt and the largest insertion detected is 111 nt. 61.1% of deletions are less than 11 nt with the longest deletion being 87 nt. Together, only about 1% of detected INDELs over nt 1800–1840 region were larger than 50 bp.

**Table 1.** The size distribution of INDELs detected in dsl-cccDNA over nt 1800–1840 in patients with hepatitis B infection.

| Type | Length (nt) |  |  |  |  |  |
| --- | --- | --- | --- | --- | --- | --- |
|  | 1–10 | 11–20 | 21–30 | 31–40 | 41–50 | 50+, Max (bp) |
| Insertion | 80.8% | 11.8% | 3.2% | 1.6% | 1.4% | 1.1%, 111 |
| Deletion | 61.6% | 17.0% | 11.4% | 4.2% | 4.8% | 1.0%, 87 |

### Prevalence of dsl-cccDNA

As we concluded that deletions clustered in the nt 1750 region can be found in authentic cccDNA, we therefore define dsl-cccDNA by only the INDELs found at the DR1 end-joining site (nt 1800–1840). Next, we determined the prevalence of dsl-cccDNA in this study cohort. Of the 32 subjects, the INDEL at the DR1 end-joining site was detected in all 32 subjects, indicating a 100% prevalence of dsl-cccDNA for patients that contain detectable cccDNA. We next calculated the percentage (%) of dsl-cccDNA of total cccDNA, as summarized in **Figure 5**. Although dsl-cccDNA was detectable in all patients having detectable cccDNA, the majority of cccDNA detected (71.8–97.8%) are authentic cccDNA without detectable insertion or deletions as indicated in grey (wild-type, WT). Of detected INDELs, the templates with insertions only (0.76–26.4%) are significantly (p<0.001 by Wilcox Signed Rank test) more abundant followed by the templates with deletions only (0.08–3.00%). Only a small fraction (0.0004–1.73%) of the templates contain both deletions and insertions. Of 32 CHB patients with detectable cccDNA, 18 (56%) contained less than 10% of dsl-cccDNA. Only 2 (6%) had over 20% of cccDNA detected that were dsl-cccDNA as summarized in **Figure 5B**.

**Figure 5.**
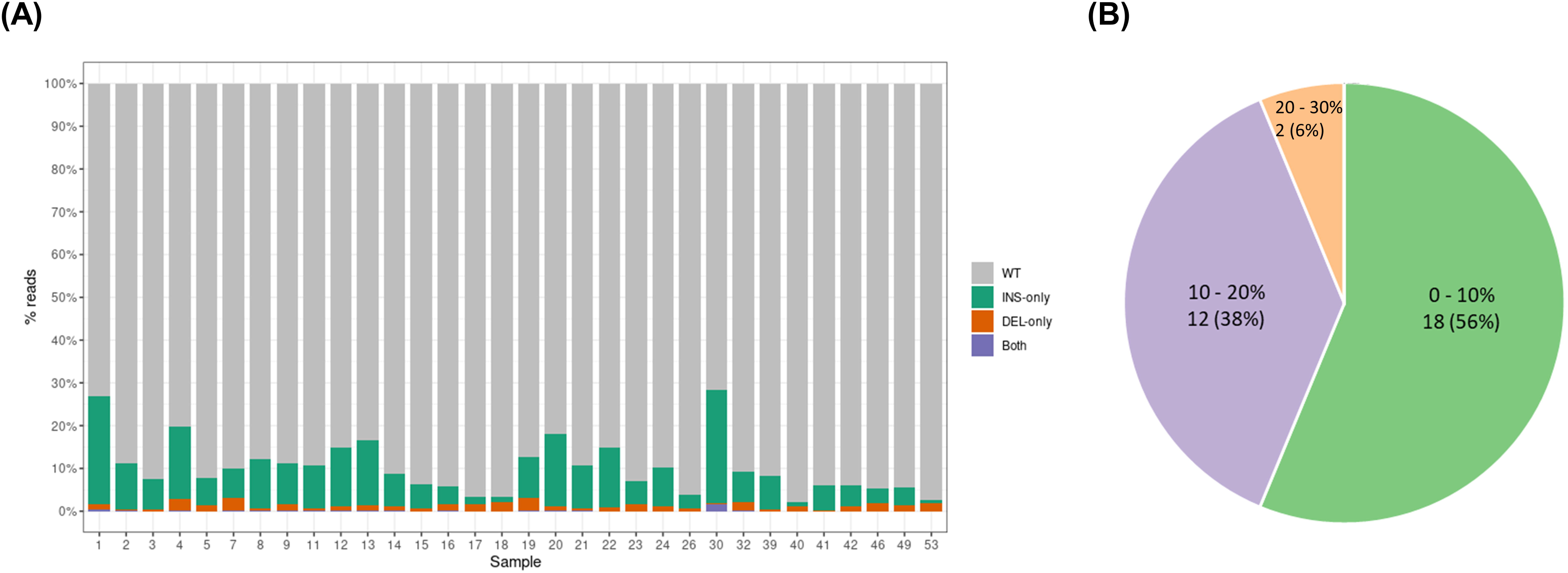
Prevalence of intrahepatic dsl-cccDNA (INDELs occurring in nt 1800–1840) in the CHB patients as determined by PSAD-cccDNA PCR-NGS assay. (A) The compositions of intrahepatic cccDNA and dsl-cccDNA for each patient. (B) The overall presence of dsl-cccDNA in the cccDNA detected in 32 CHB patients.

### Dsl-cccDNA is more abundant in HBeAg(+) CHB patients

As dsl-cccDNA is a by-product of HBV replication, it was of interest to determine if dsl-cccDNA is in higher abundance in HBeAg(+) samples. As its precursor, dsl-DNA should be more abundant in these HBeAg(+) CHB patients. As shown in **Figure 6A**, the proportion of dsl-cccDNA in total cccDNA was higher in HBeAg(+) group compared to HBeAg(-) group with mean 11.3 (3.2–26.9)% *vs.* 7.7 (2.2–28.2)% (p=0.01). There was no significant correlation between dsl-cccDNA and cccDNA levels (r=0.08, p=0.65]. Next, we determined the diversity of dsl-cccDNA species and its correlation with various viral factors, such as cccDNA level and other viral blood biomarkers. The diversity of dsl-cccDNA species, as one example shown in **Figure 2B**, was calculated by Shannon’s diversity index as detailed in Materials and Methods. As shown in **Figure 6B**, the dsl-cccDNA diversity significantly correlates with cccDNA levels (log-transformed; r=0.82, p<0.001), and is different between HBeAg levels (p<0.001) and serum HBV DNA loads (p<0.001).

**Figure 6.**
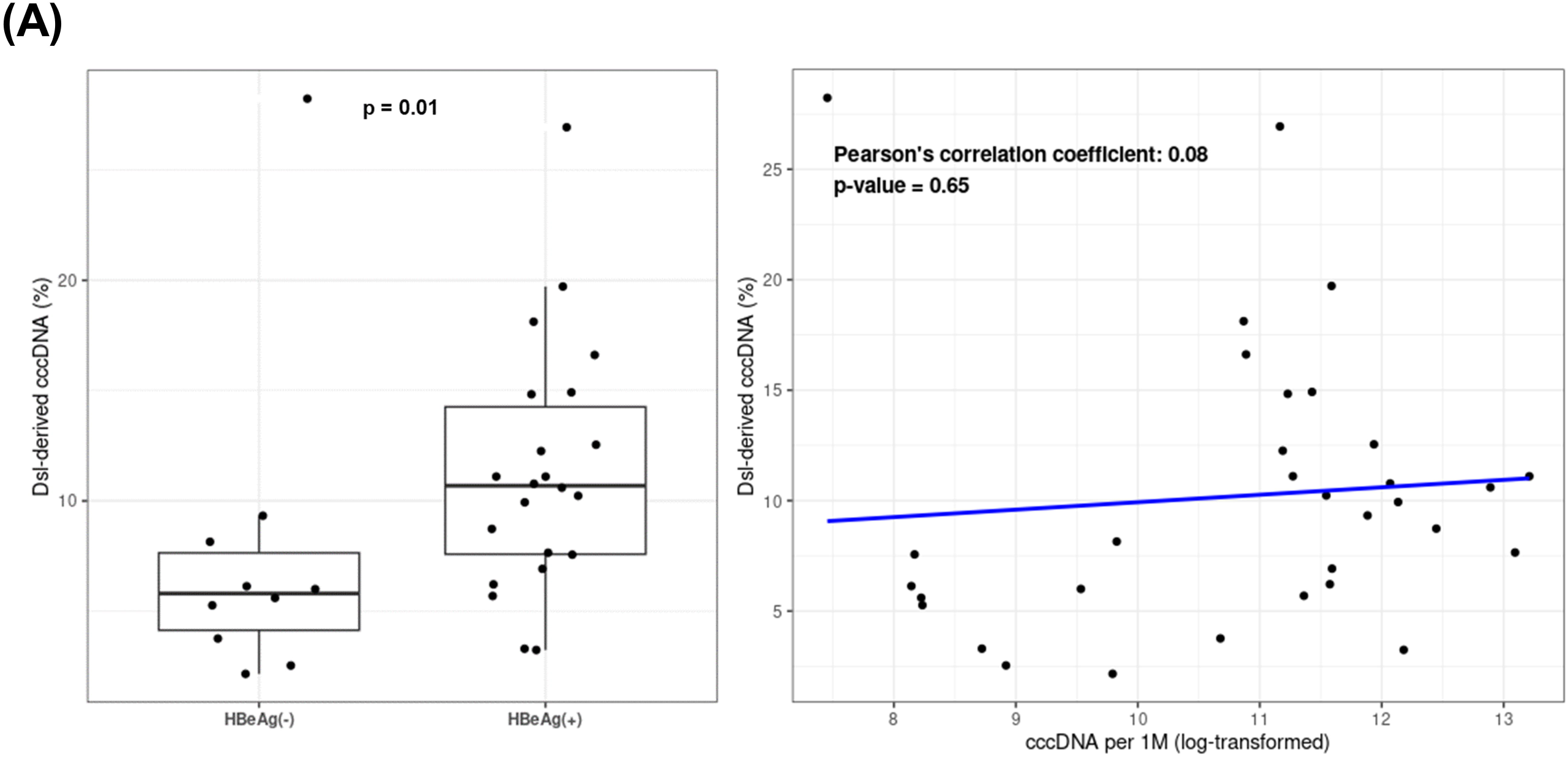
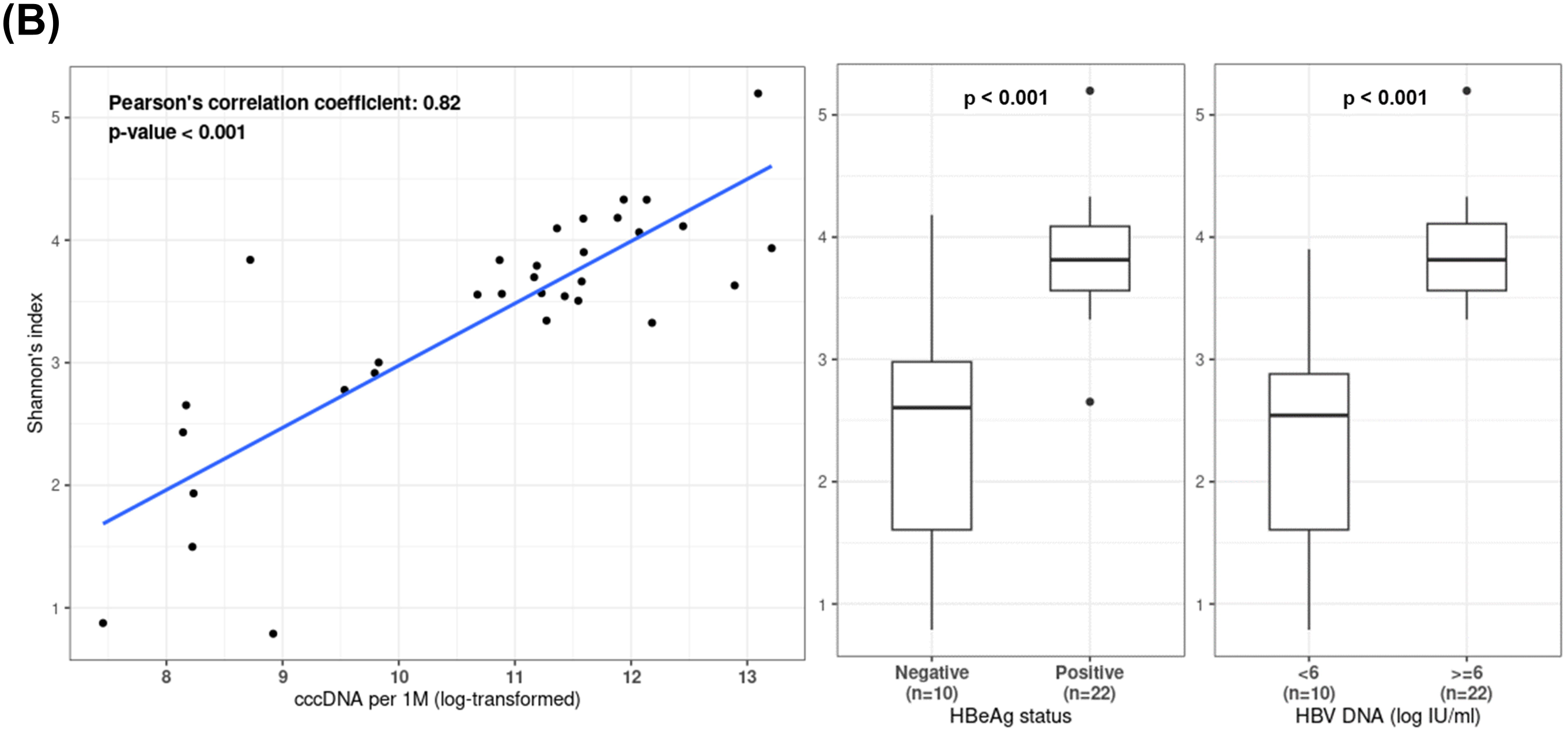
Characterization of dsl-cccDNA with other viral factors. (A) Comparison of the prevalence of dsl-cccDNA with HBeAg status (left; p=0.01; Wilcoxon’s Rank Sum test) and its correlation with the cccDNA level (right; p=0.65; Pearson’s correlation test). (B) Relationships of the diversity of dsl-cccDNA with the level of cccDNA (left; p<0.001; Pearson’s correlation test), HBeAg (middle; p<0.001; Wilcoxon’s Rank Sum test), and serum HBV DNA load (right; p<0.001; Wilcoxon’s Rank Sum test).

## Discussion

The hepadnaviral dsl-cccDNA has been shown in cell cultures and animal (duck and woodchuck) models for over 2 decades, this study is the first report that detected and characterized dsl-cccDNA in livers from CHB patients. By a concordance study from two independent NGS studies, HBV-targeted NGS of 56 liver needle biopsies and PSAD-cccDNA-PCR NGS of 32 of 56 samples, we characterized the dsl-cccDNA species by the positions and sizes of INDELs around the DR1 site centered at nt 1820. A 22-nt deletion in the *X*-gene was discovered in cccDNA in all 32 studied patients by PSAD-cccDNA-PCR NGS assay (**Figure 4**). By comparing the percentage of dsl-cccDNA in total cccDNA between the 22 HBeAg(+) and 10 HBeAg(-), the dsl-cccDNA was significantly more frequent among samples with HBeAg(+) (p=0.01), independent of the levels of their viremia (**Figure 6A**). Interestingly, the diversity of dsl-cccDNA species correlates with cccDNA levels (r=0.82, p<0.001), and is different between HBeAg levels (p<0.001) and serum HBV DNA loads (p<0.001) (**Figure 6B**).

According to the nature of dsl-cccDNA formation *via* the error-prone NHEJ machinery, different self-ligation events likely result in various joint sequences with different insertions or deletions, thus INDELs in dsl-cccDNA should only be supported by 1-few SRs. This study first identified samples that likely to contain dsl-cccDNA by detection of the “dsl-cccDNA-like” INDELs at DR1 end joint site (nt 1800–1840), as an example shown in **Figure 2B** by HBV-targeted NGS assay of 56 CHB tissue biopies. As outlined in **Figure 1**, we selected 32 samples, including two negative controls, for PSAD-cccDNA-PCR NGS assay for the concordance study to identify and characterize dsl-cccDNA in infected livers.

Four observations are documented in this study. **First**, we detected a high prevalence of dsl-cccDNA in CHB patients in our study cohort. All 32 liver biopsies contained detectable dsl-cccDNA by PSAD-cccDNA-PCR NGS assay including the two negative controls selected by HBV-targeted NGS assay. This is not unexpected since cccDNA-PCR NGS assay focusing on the DR2-1 region providing at least 100-times more resolution, thus more sensitive to detect INDELs than HBV-targeted NGS assay and self-ligation of dslDNA forming dsl-cccDNA could be as frequent as recomination with host DNA resulting in integration which is high prevalence. **Second,** in consistent with previous reports from tissue culture and animal studies [9, 10] that we detected heterogenous pattern of INDELs. We did not detect any INDEL species at the DR1 joint site (nt 1800–1840) having over 10 SRs. The dsl-cccDNA in CHB patients should be mostly defective, if any for further DNA synthesis, may be only few generations. Thus, the profile of dsl-cccDNA pool could represent a mixture of cccDNA converted from the infecting viruses carrying dslDNA genomes and progeny dsl-cccDNA replenished by illegitimate replication, plus those converted from dslDNA byproduct during legitimate replication supported by authentic cccDNA. This pool of INDELs are mediated by error-prone NHEJ DNA repair pathway, giving rise to an extensive and heterogenous pattern of INDELs at DR1 region. **Third,** most of dsl-cccDNA INDELs are shorter than 10 nt with the largest deletion and insertion detected spanning 111 bp and 87 bp, respectively (**Table 1, Supplemental Figure 5**). Through an analysis of deletion sizes, we discerned that the most substantial deletion observed was 87 bp, with deletions larger than 50 bp constituting a mere 1% of the total. An overwhelming 90% of deletions were limited to 30 bp or less.

**Fourth**, the discovery of the 22-nt deletion (nt 1755–1776) that may impact X/BCP/ENII sequence that was validated by PCR cloning and Sanger sequencing of tissue biopsy DNA (**Supplemental Figure 1**), was found universally presented in all 32 study CHB subjects. By further sequence analysis using BLAST of 119-nt WT and 97-nt deleted sequences to the deposited sequences in GenBank, a range of identity from 80% – 100% was shown in the 100 most homologous complete genome hits (**Supplemental Table 3**). Interestingly, although we did not find an identical 22-nt deletion sequence in the GenBank, 78 of these 100 most highly homologous hits sequences have at least 90% identity to this 22-nt deleted 97-nt sequences. We thus conclude this 22-nt deletion is in the similar region of previously reported 20-nt heterogeneous deletions, nt 1753*–*1777, reported with dysfunctional X-protein, as part of HBV quasi-species [20]. Interestingly, this study [20] also showed that INDEL mutations were more prevalent in HBeAg-positive CHB patients prior to antiviral treatment and a higher detection rate of these mutations at baseline might correlate with a better response to LMV and ADV combination therapy. Consistent with these previously reported 20-nt deletions in this region, this 22-nt deletion is in a range of <1% to 46% of cccDNA, determined by cccDNA-PCR NGS study (data not shown). These 20–22-nt heterogeneous deletions in X-gene may be developed as a potential biomarker for treatment response.

Although this study represents the first comprehensive characterization of dsl-cccDNA and its prevalence among CHB patients, it is important to note that all the dsl-cccDNA reported are limited in the cccDNA species containing PCR amplicon sequences, nt 1553–1949. It is likely that other noncanonical shorter dslDNA forms may arise during the viral replication, indicating the potential existence of other smaller-sized dsl-cccDNA in infected liver. Further investigations are warranted to elucidate the presence of other dsl-cccDNA species.

As dsl-cccDNA is a self-ligation of dslDNA, byproducts during viral replication, we hypothesized that more active viral replication should generate more copies (quantity) and more variants (diversity) of dsl-cccDNA. As expected, its quantity and diversity are significantly higher in patients with active viral replication as indicated by HBeAg status (**Figure 6A and 6B, respectively**) and HBV DNA levels (**Figure 6B**), which is in line with our hypothesis. It is worth noting that, similar to the integrated HBV DNA [21], the majority of dsl-cccDNA possess the promoters and ORFs of HBV surface antigens and may be competent for HBsAg expression if the HBV polyadenylation signal downstream of DR1 remains intact and functional, however, the loss of HBeAg and HBx coding capacity due to ORF disruption by INDELs, especially the latter, would require the presence of authentic cccDNA in the same cell to provide HBx *in trans* to activate dsl-cccDNA episome transcription. Chronic hepatitis in humans is known to be associated with the progressive appearance and accumulation of hepatocytes that display low or undetectable levels of virus antigen expression or abnormal patterns of expression. We suggest that pleomorphic virus expression in infected livers may be correlated with the presence of hepatocytes that express defective cccDNA molecules. More studies are needed to further investigate the role and regulation of intrahepatic dsl-cccDNA in CHB infection and development of HCC.

## Abbreviations (alphabetical order)

chronic hepatitis B (CHB), double-stranded linear DNA (dslDNA), dslDNA derived covalently closed circular DNAs (dsl-cccDNAs), Hepatitis B Research Network (HBRN), insertions and deletions (INDELs), next-generation sequencing (NGS), nonhomologous end-joining (NHEJ), plasmid-safe ATP-dependent (PSAD), relaxed circular DNA (rcDNA), supporting reads (SRs).

## Data Availability

All data produced in the present study are available upon reasonable request to the authors.

## Acknowledgments

This study is supported by grants from the National Institutes of Health (R56AI179574 to DL, YS, and HG; R01AI110762, R01AI150255, R21AI179929 to HG; U01DK082919-010 to DL; U01CA275648 to YS; R43AI167169 to SL); R01AI155140 to RTC.

