## Supplemental Table 1, Supplemental Figure 1, 2, 4, 5 for "Detection and characterization of Hepatitis B virus double-stranded linear DNA-derived covalently closed circular DNA in chronic hepatitis B patients"

**Supplemental Table 1. Baseline demographics and clinical features.**

|  | **HBeAg(+)**  **n=24** | **HBeAg(-)**  **n=32** | **P value** |
| --- | --- | --- | --- |
| **Age (years)** | 42.6  (23–66.6) | 46.5  (21.9–68.9) | 0.09 |
| **Gender (M:F)** | 12:12 | 22:10 |  |
| **Race**  **Asian**  **White**  **Black**  **Other** | 17  4  2  1 | 25  3  3  1 |  |
| **ALT (ULN)** | 3.9  (0.8–11.4) | 2.9  (0.8–21) |  |
| **Serum HBV DNA (log_10_ IU/ml)** | 7.5  (3.2–9.2) | 4.5  (2.6–7.7) | <0.00001 |
| **Serum qHBsAg (log_10_ IU/ml)** | 4.25  (1.94–5.35) | 2.96  (0.5–4.15) | <0.00001 |
| **HBV genotype**  **A**  **B**  **C**  **D** | 5  4  11  4 | 5  15  7  5 |  |

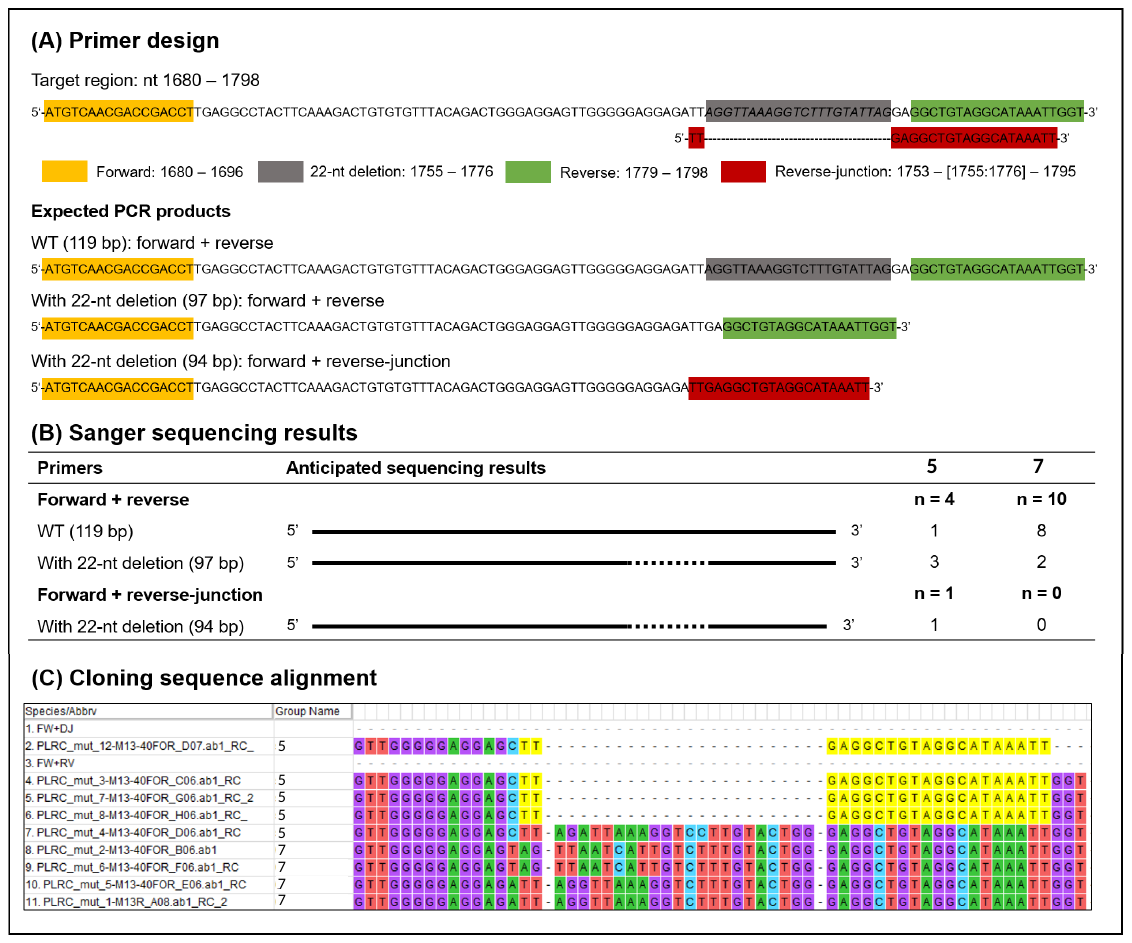

**Supplemental Figure 1**. (A) Primer design for 22-nt deletion validation. One forward and two reverse primers were picked to make two sets of primers and three expected products. The reverse primer could amplify both sequences in WT (110 bp) and with 22-nt deletion (97 bp), while the reverse deletion-specific primer can only amplify sequences with this deletion (94 bp). (B) Sanger sequencing results of PCR cloning of the three types of products. (C) Cloning sequence alignment at the 22-nt deletion site.

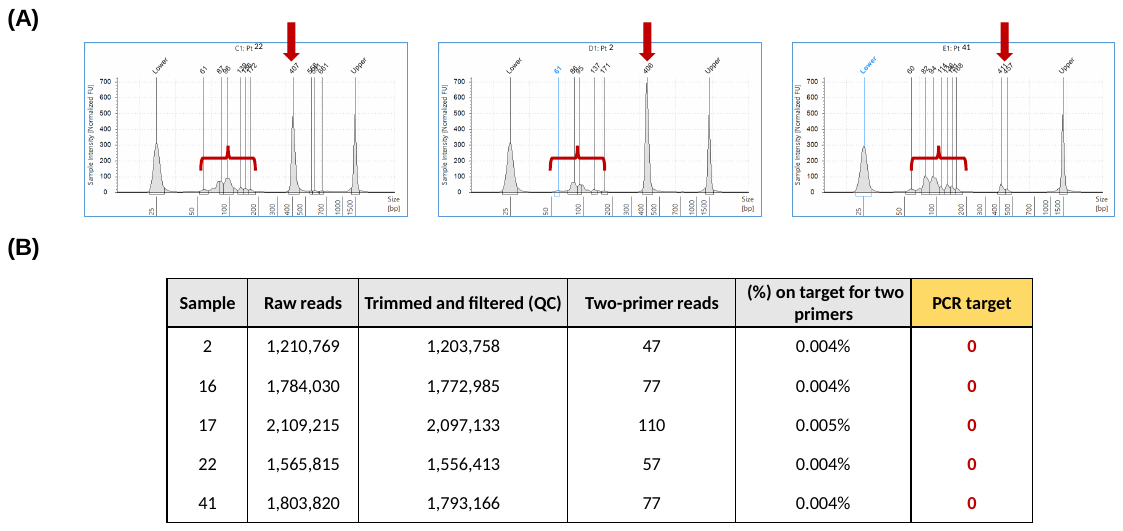

**Supplemental Figure 2.** (A) Non-specific short (50–200 bp) PCR products from PSAD cccDNA assay revealed by Tapestation capillary electrophoresis. The arrows pointed to the position of the anticipated full-length PCR product of 397 bp. The short PCR products in a range of 50–200 bp were also noted. (B) Summary of NGS sequencing analysis. Two-primer reads are HBV reads (reads mapped to HBV references) containing both primers regardless the entire sequences. PCR target are HBV reads that have two primers on each end with additional 30 nt anticipated HBV sequences after each primer sequences.

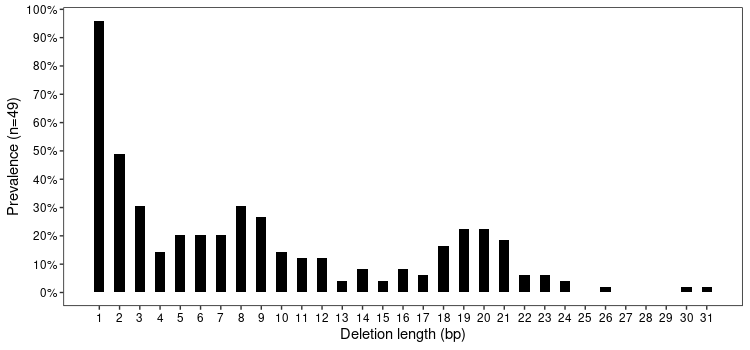

**Supplemental Figure 4**. **Deletion length distribution in nt 1750–1781.** Y-axis denotes the prevalence of each deletion length among the 49 samples that contain detectable deletions in this region.

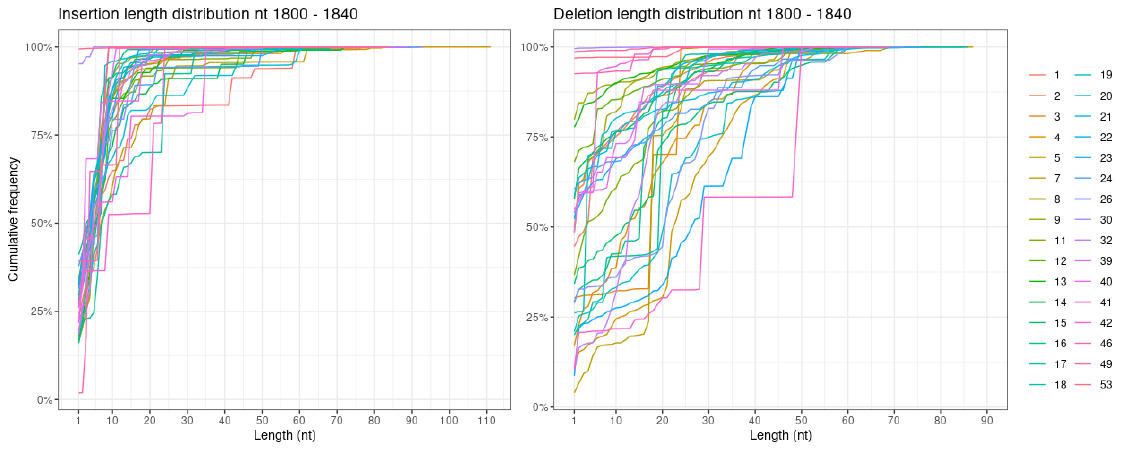

**Supplemental Figure 5**. **Cumulative frequencies of insertion and deletion lengths detected in PSAD-cccDNA PCR-NGS over nt 1800–1840 region.**
