## Supplemental Figure 3 for "Detection and characterization of Hepatitis B virus double-stranded linear DNA-derived covalently closed circular DNA in chronic hepatitis B patients"

### Slide 1
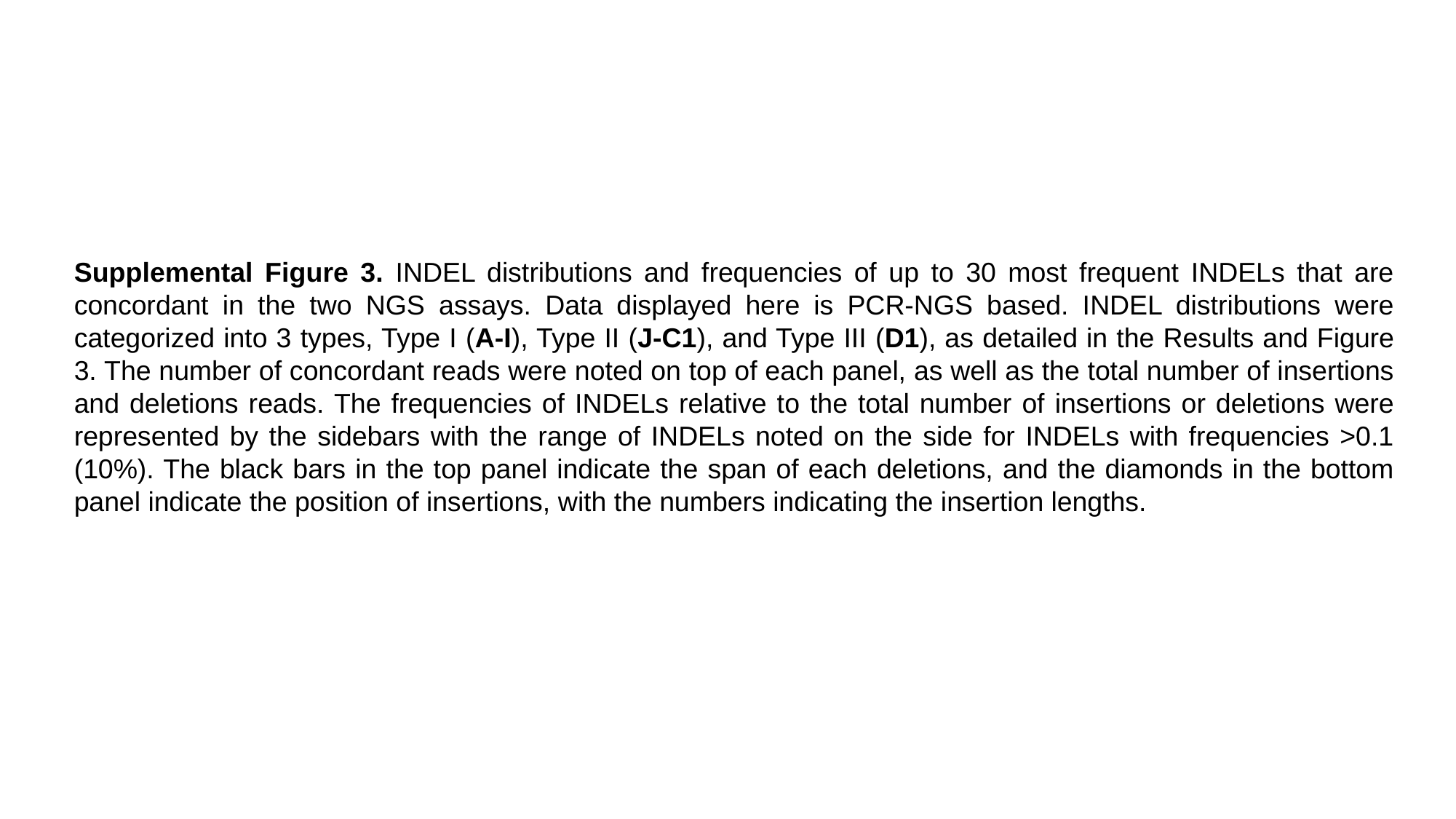

Supplemental Figure 3. INDEL distributions and frequencies of up to 30 most frequent INDELs that are concordant in the two NGS assays. Data displayed here is PCR-NGS based. INDEL distributions were categorized into 3 types, Type I (A-I), Type II (J-C1), and Type III (D1), as detailed in the Results and Figure 3. The number of concordant reads were noted on top of each panel, as well as the total number of insertions and deletions reads. The frequencies of INDELs relative to the total number of insertions or deletions were represented by the sidebars with the range of INDELs noted on the side for INDELs with frequencies >0.1 (10%). The black bars in the top panel indicate the span of each deletions, and the diamonds in the bottom panel indicate the position of insertions, with the numbers indicating the insertion lengths.

### Slide 2
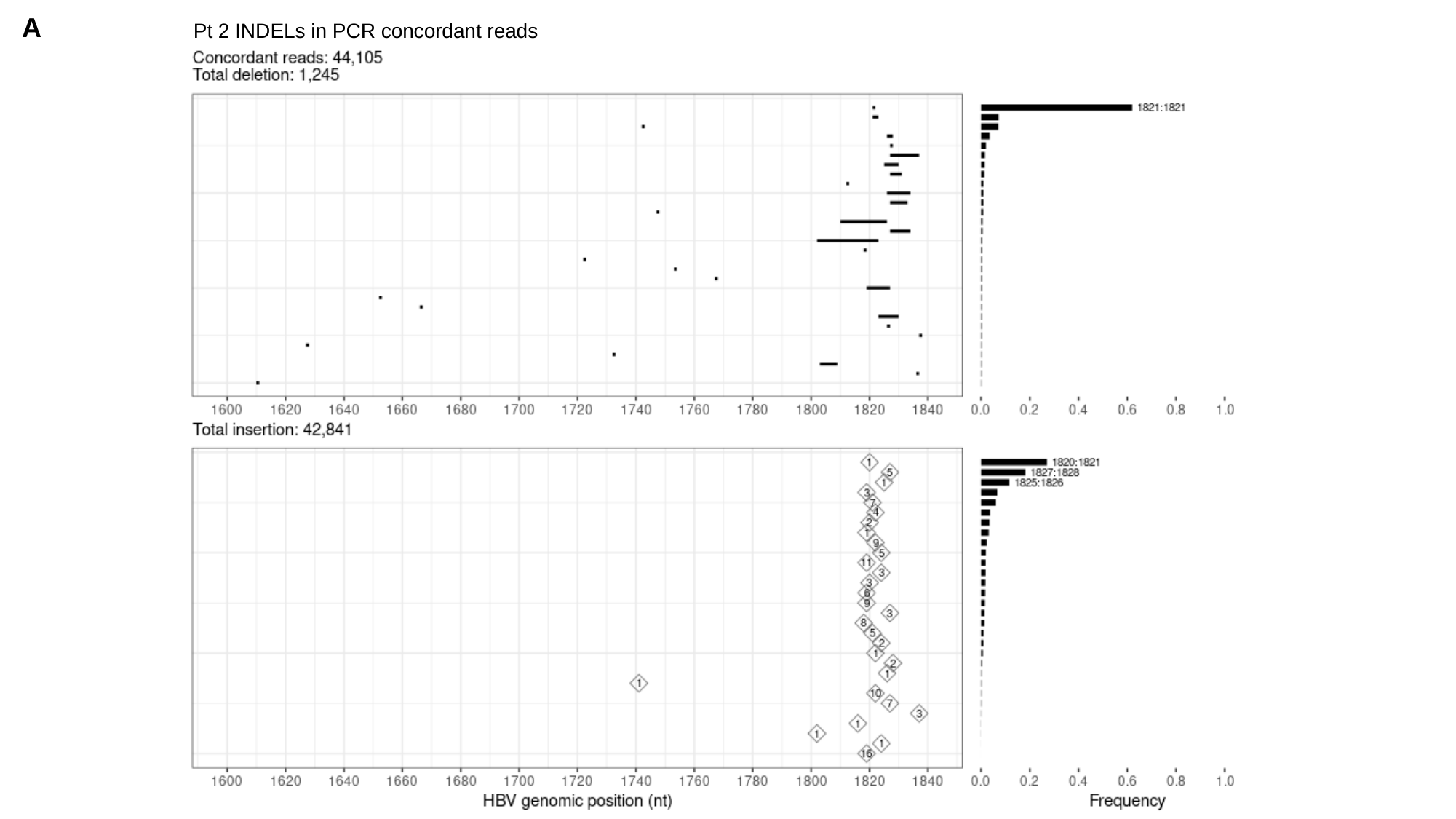

A
Pt 2 INDELs in PCR concordant reads

### Slide 3
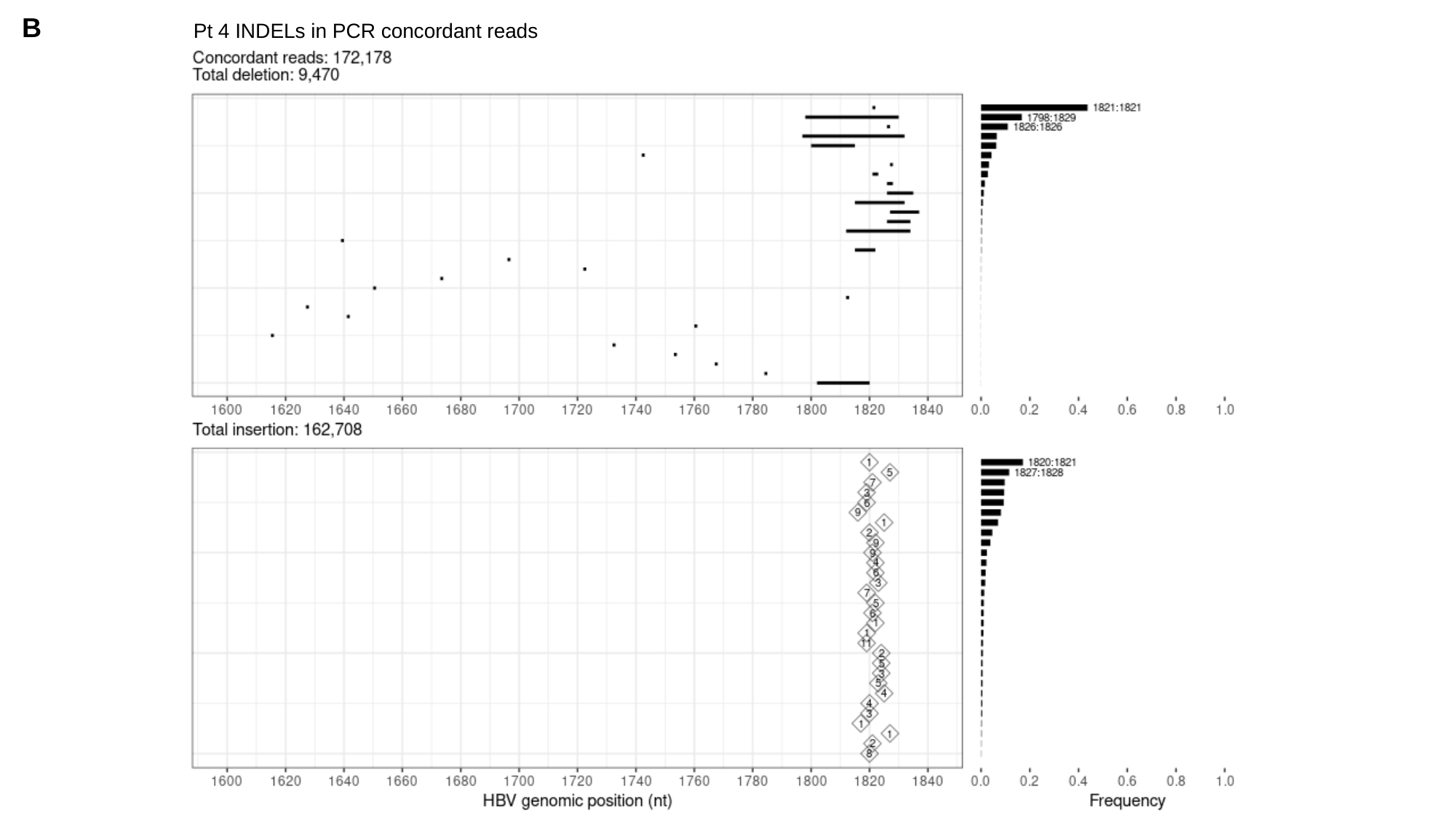

B
Pt 4 INDELs in PCR concordant reads

### Slide 4
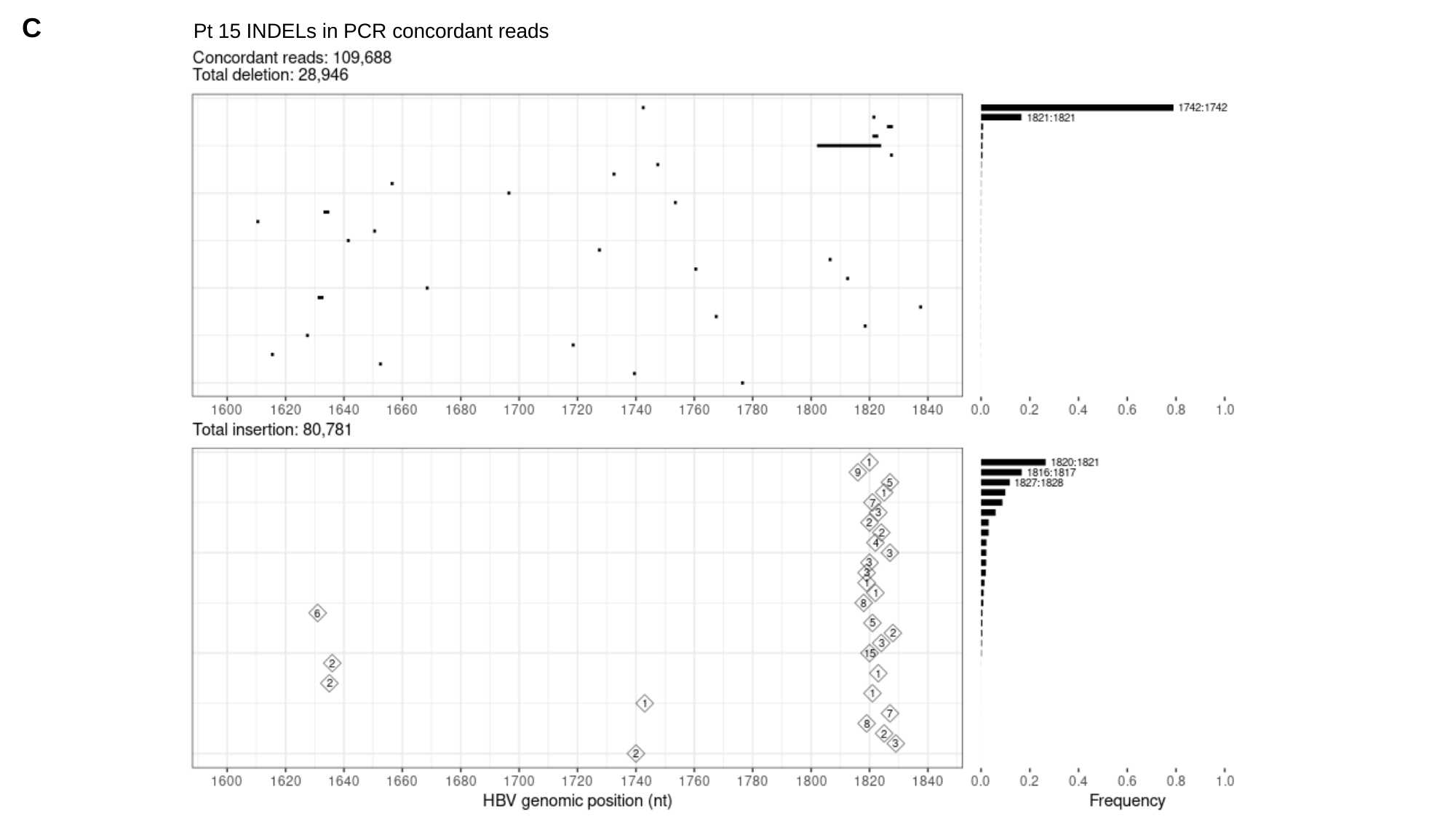

C
Pt 15 INDELs in PCR concordant reads

### Slide 5
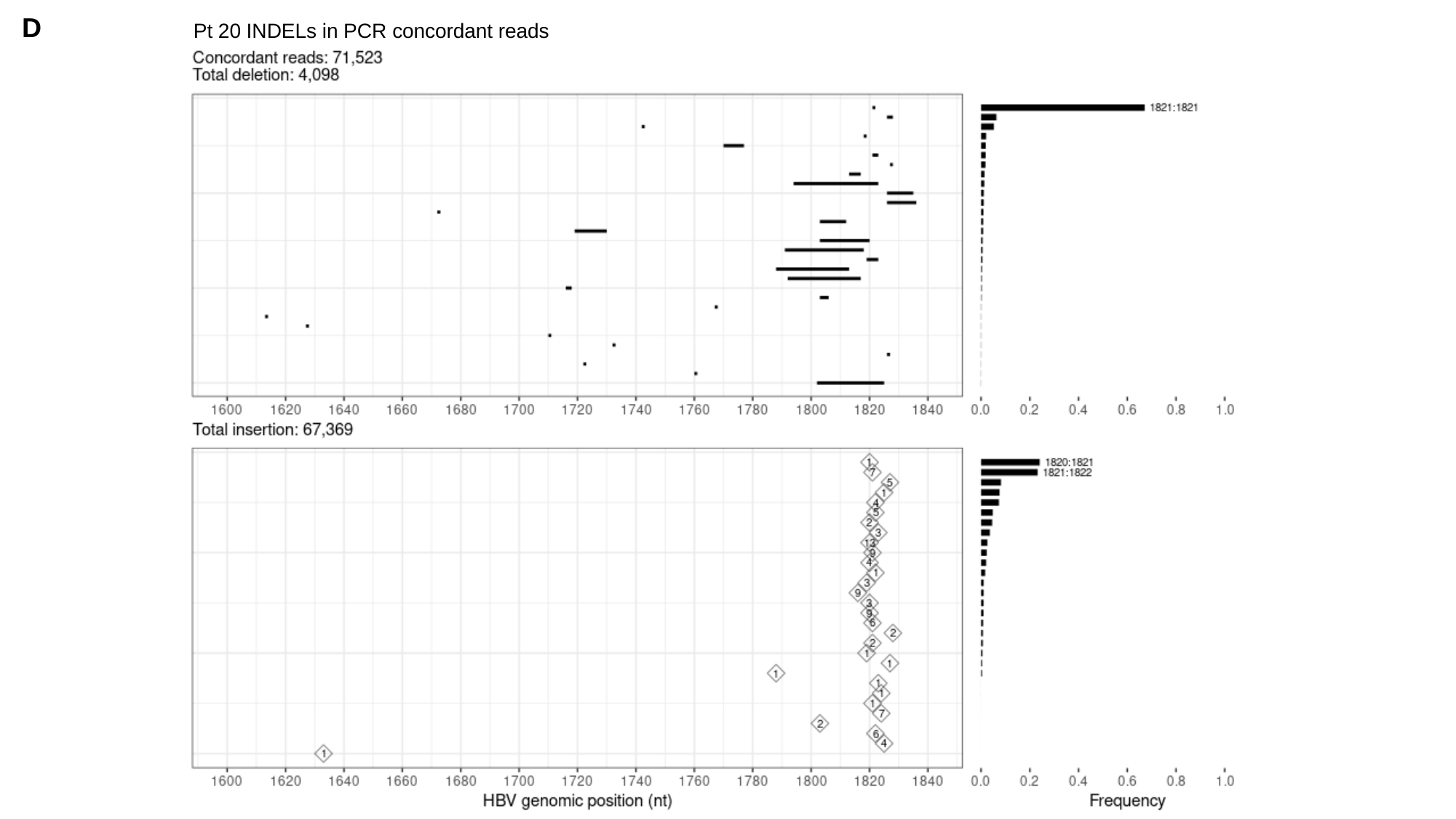

D
Pt 20 INDELs in PCR concordant reads

### Slide 6
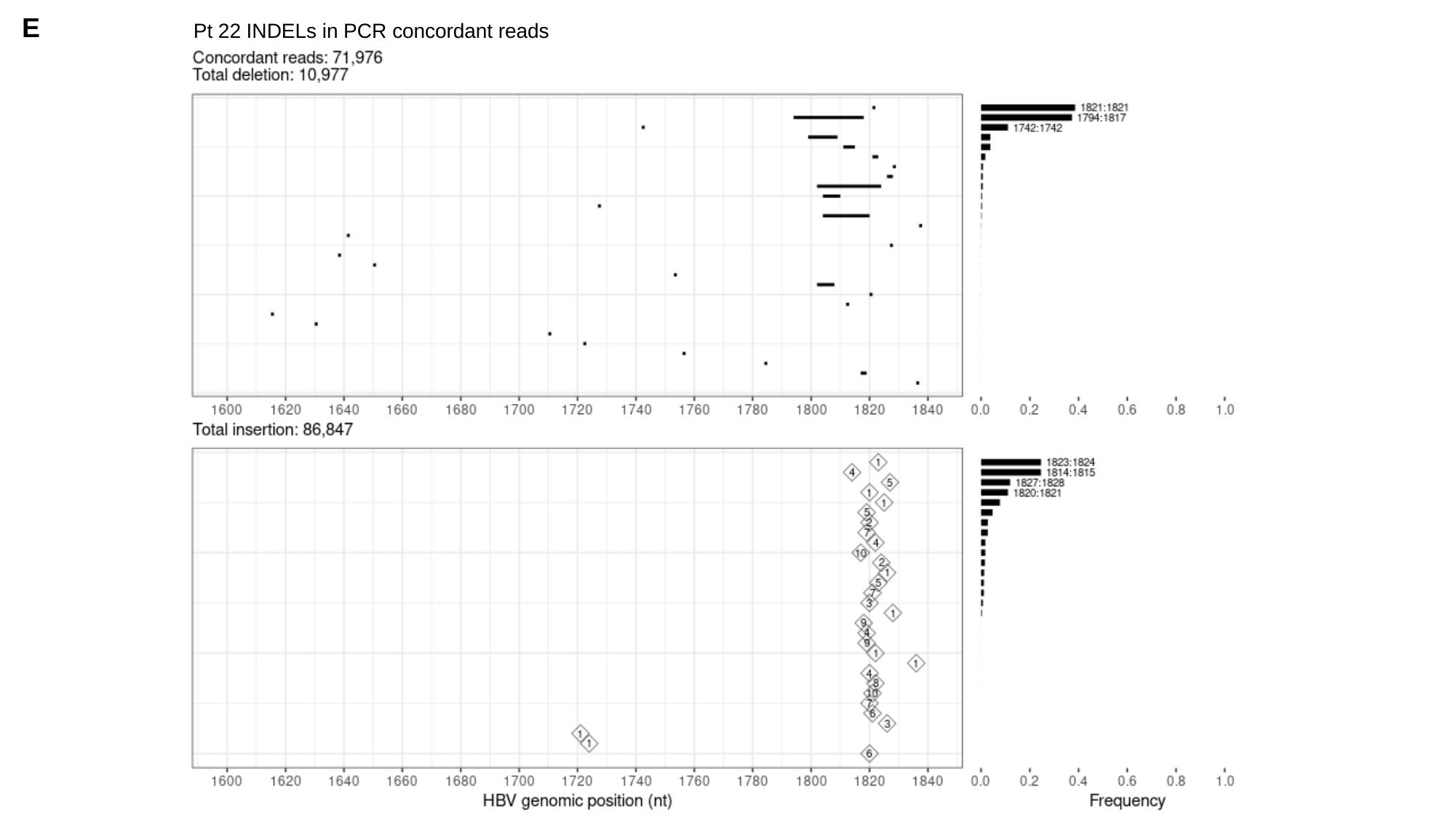

E
Pt 22 INDELs in PCR concordant reads

### Slide 7
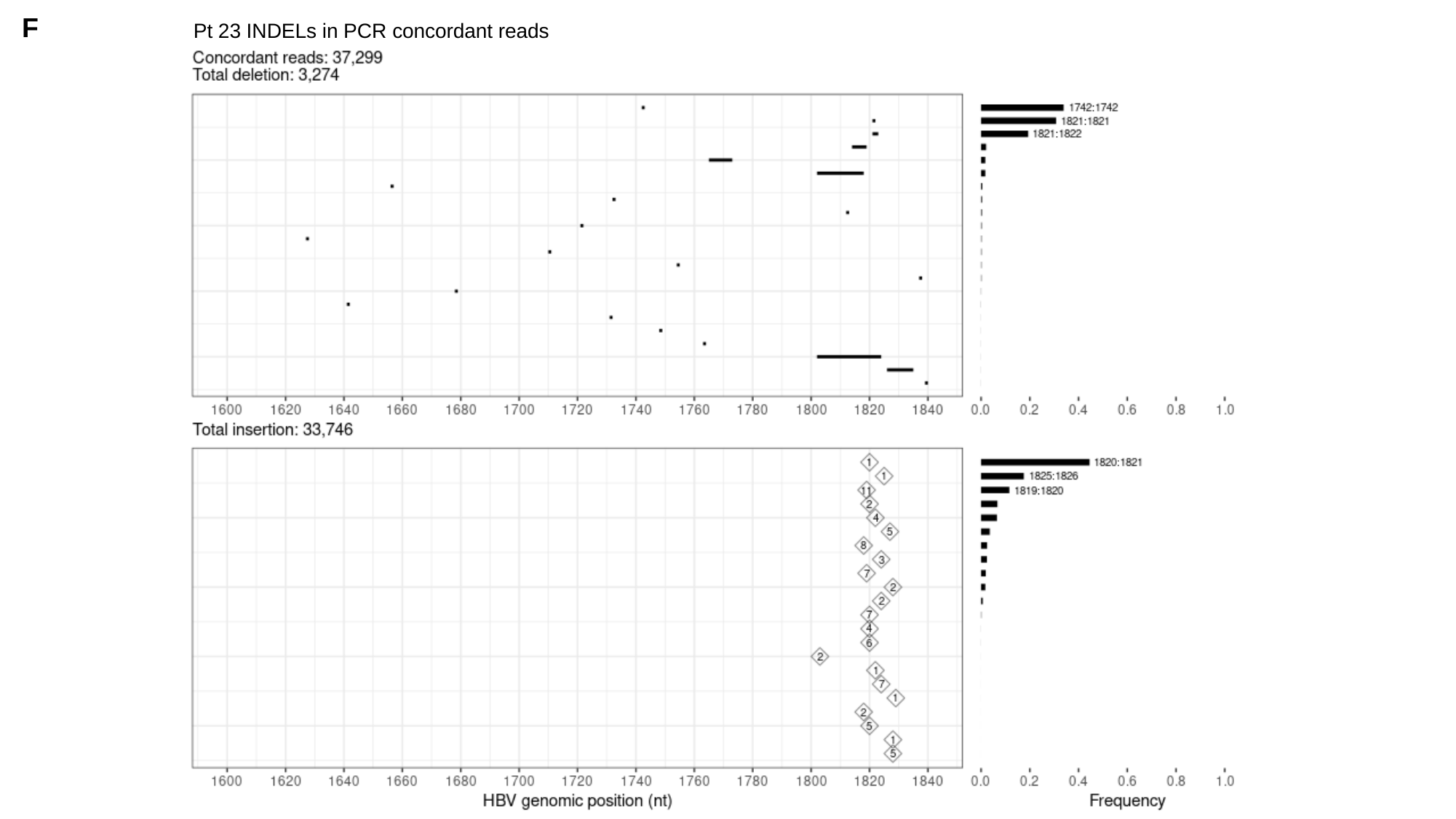

F
Pt 23 INDELs in PCR concordant reads

### Slide 8
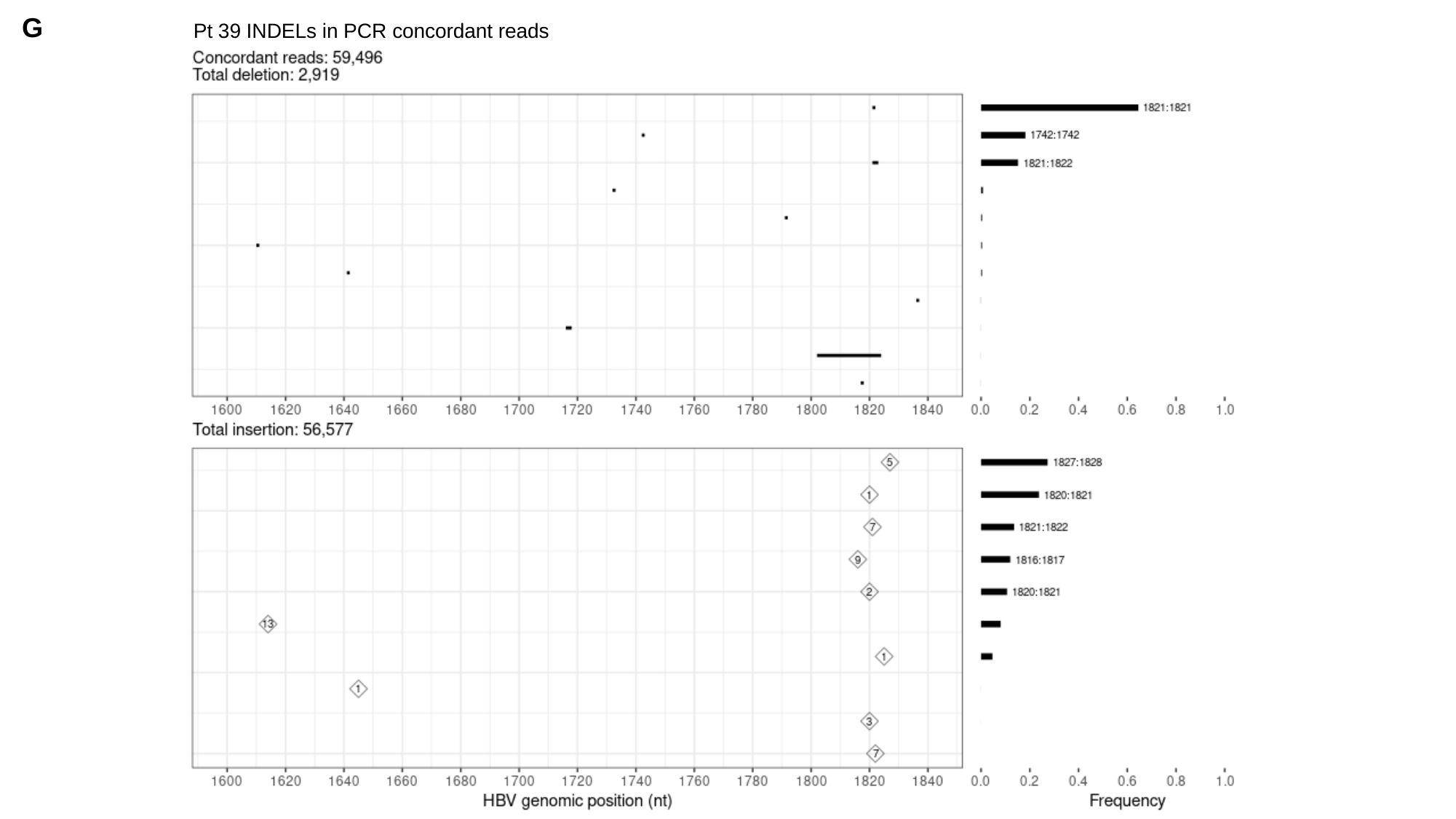

G
Pt 39 INDELs in PCR concordant reads

### Slide 9
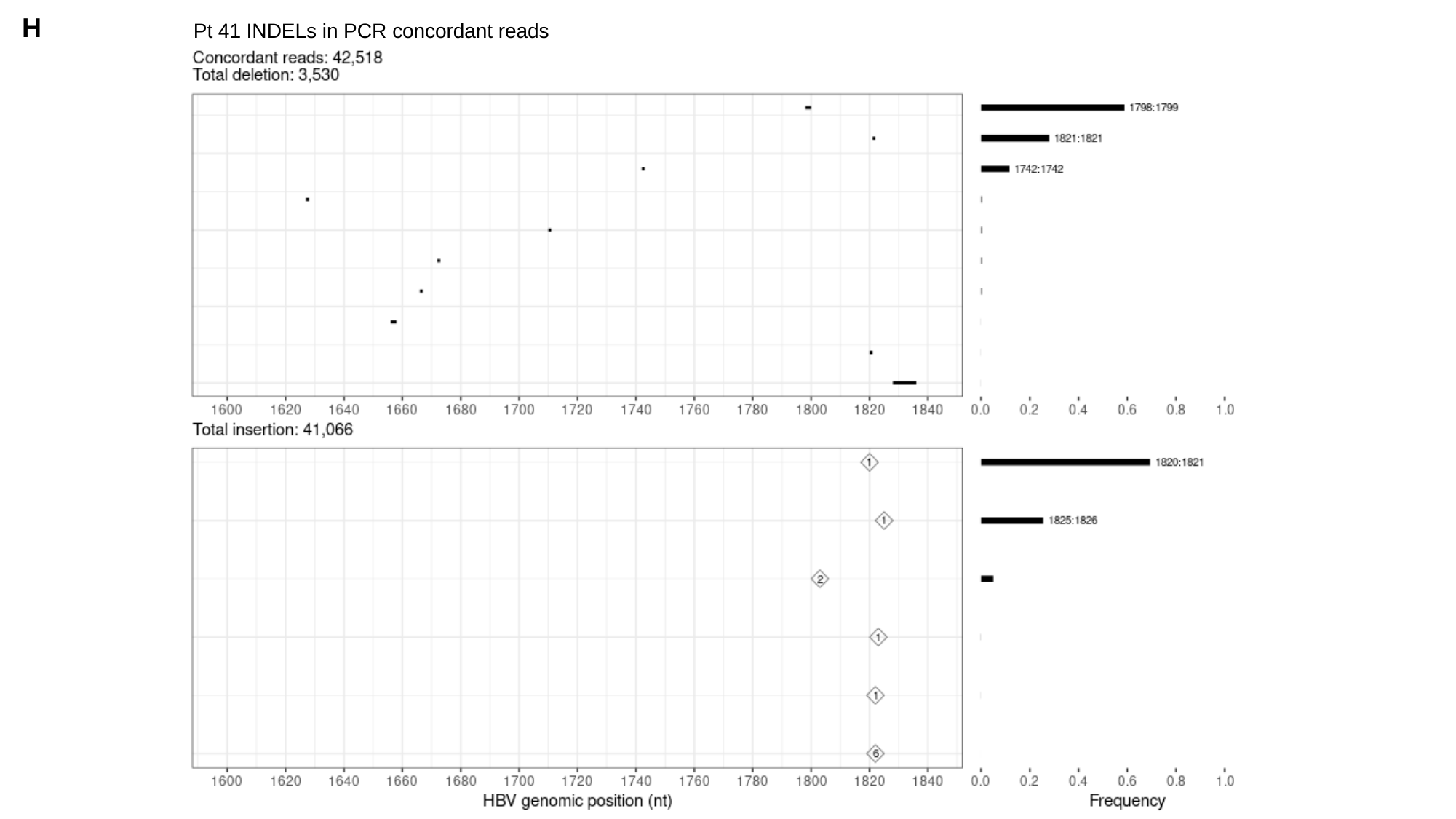

H
Pt 41 INDELs in PCR concordant reads

### Slide 10
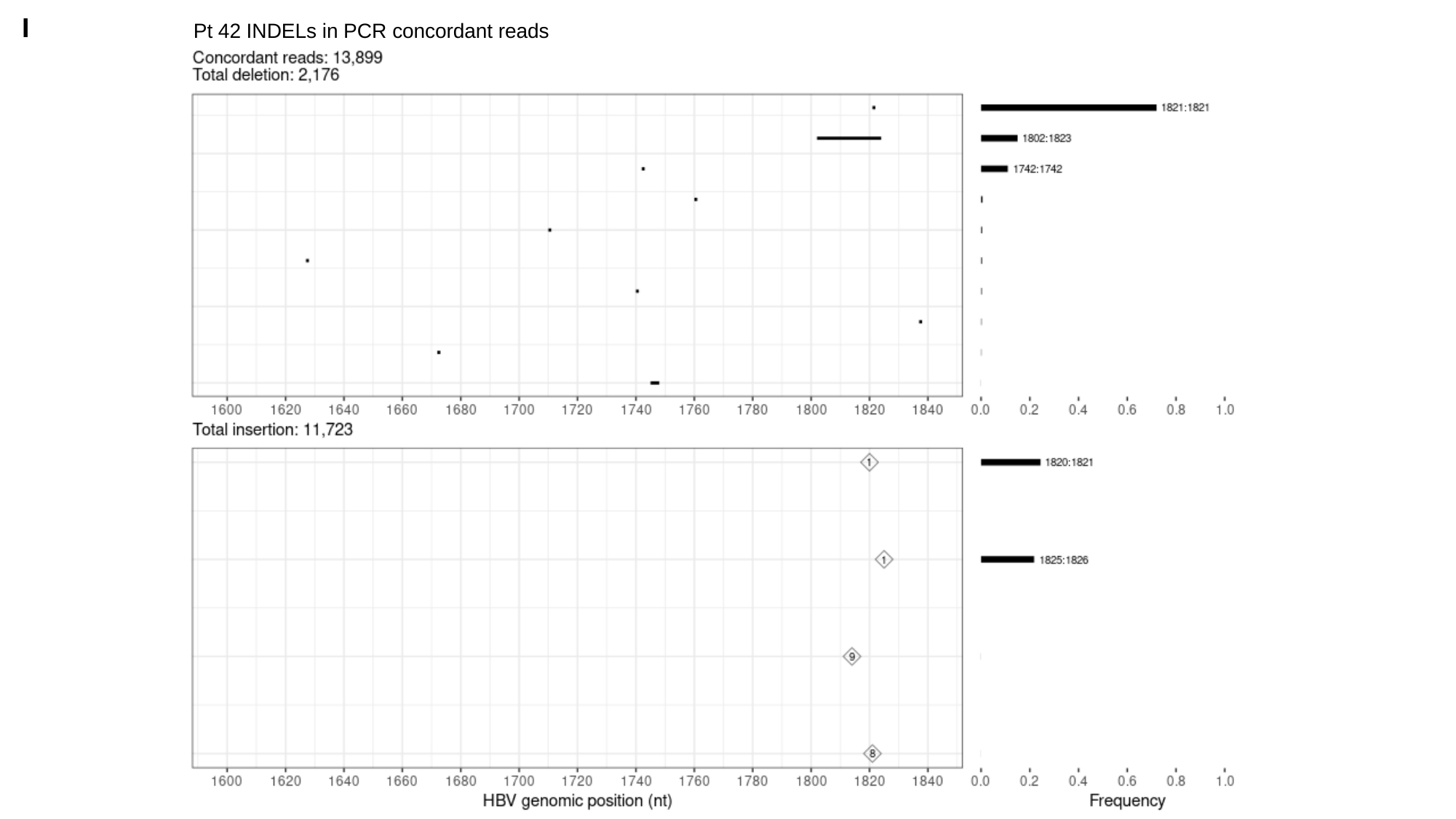

I
Pt 42 INDELs in PCR concordant reads

### Slide 11
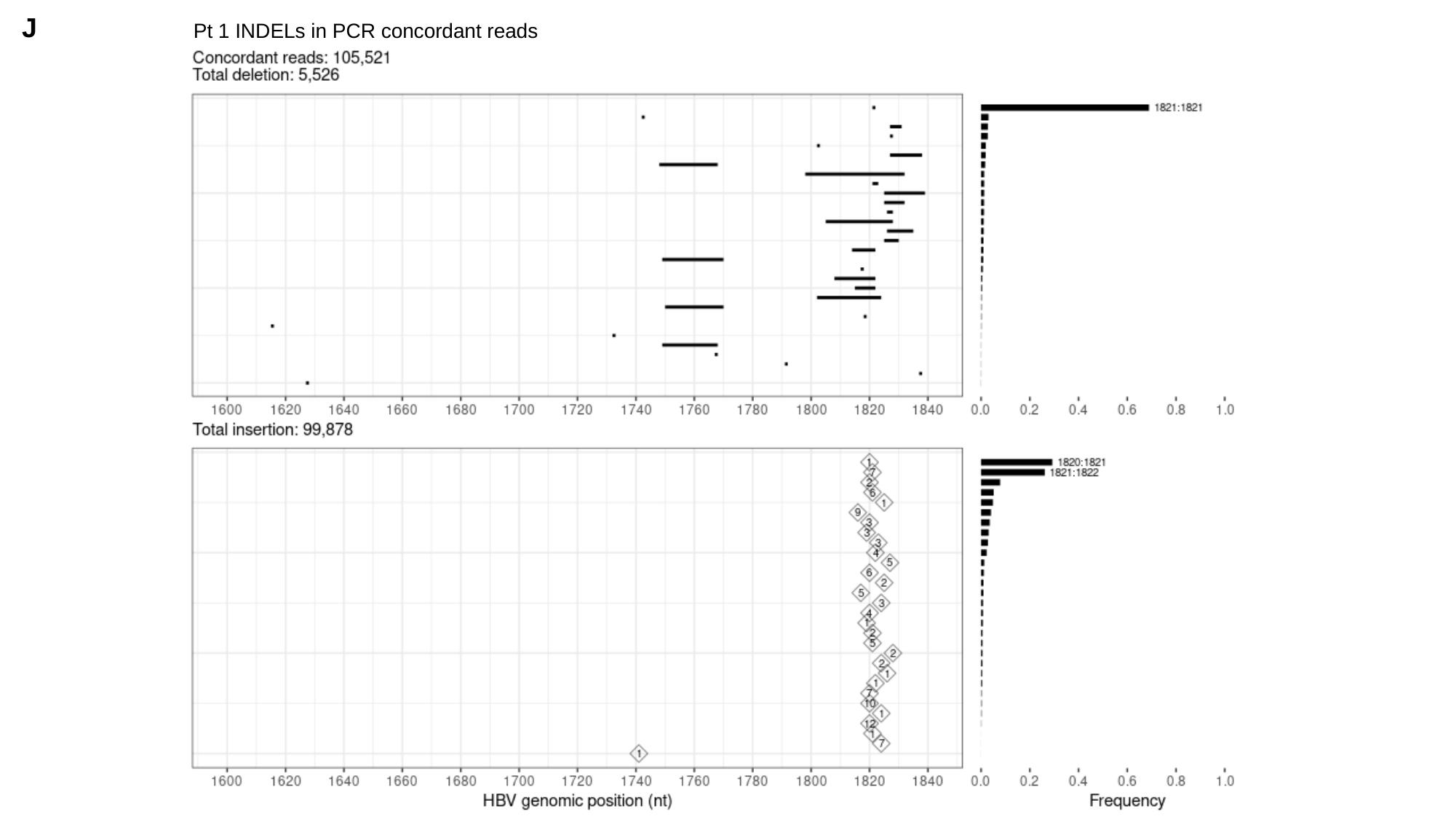

J
Pt 1 INDELs in PCR concordant reads

### Slide 12
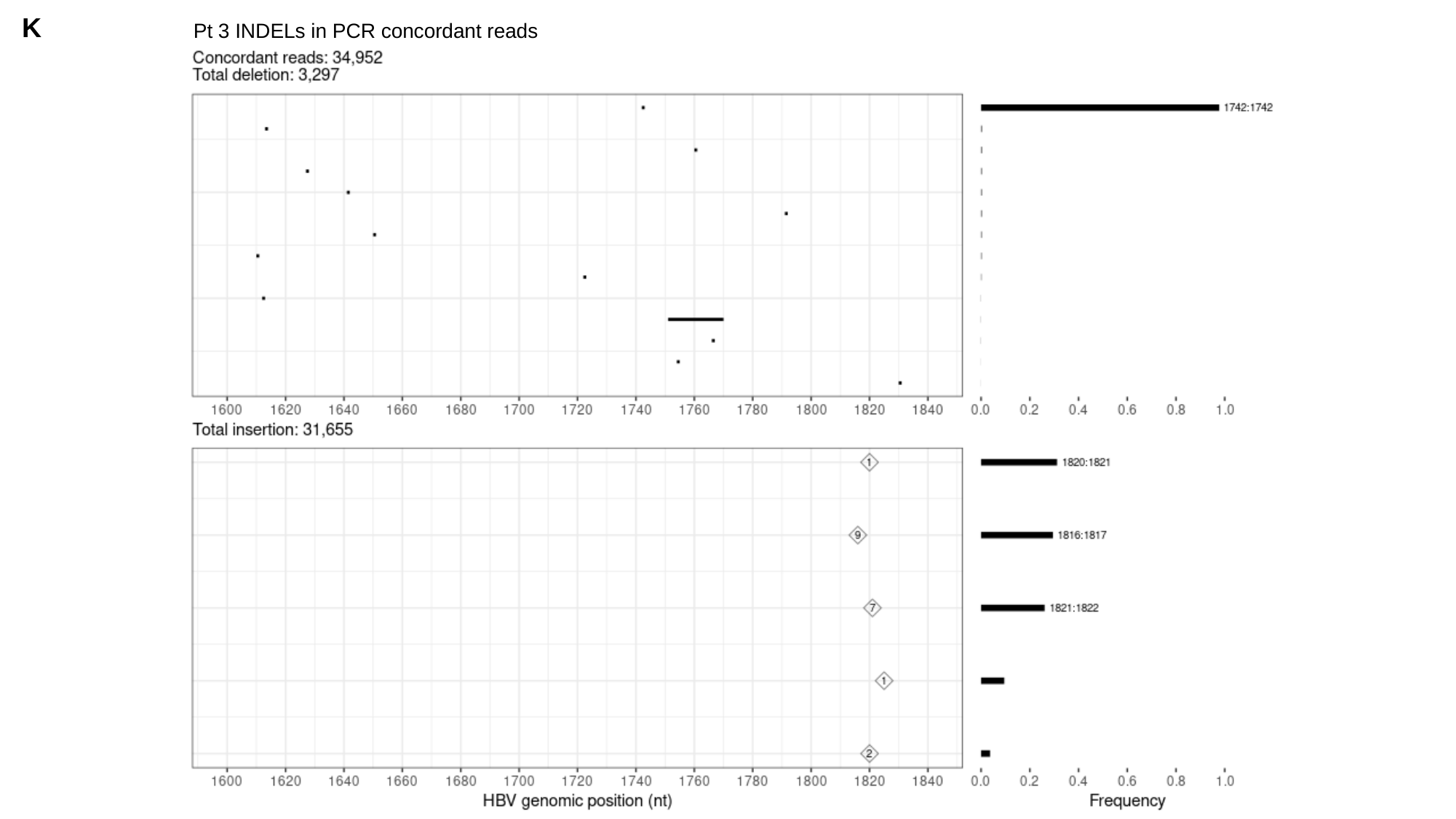

K
Pt 3 INDELs in PCR concordant reads

### Slide 13
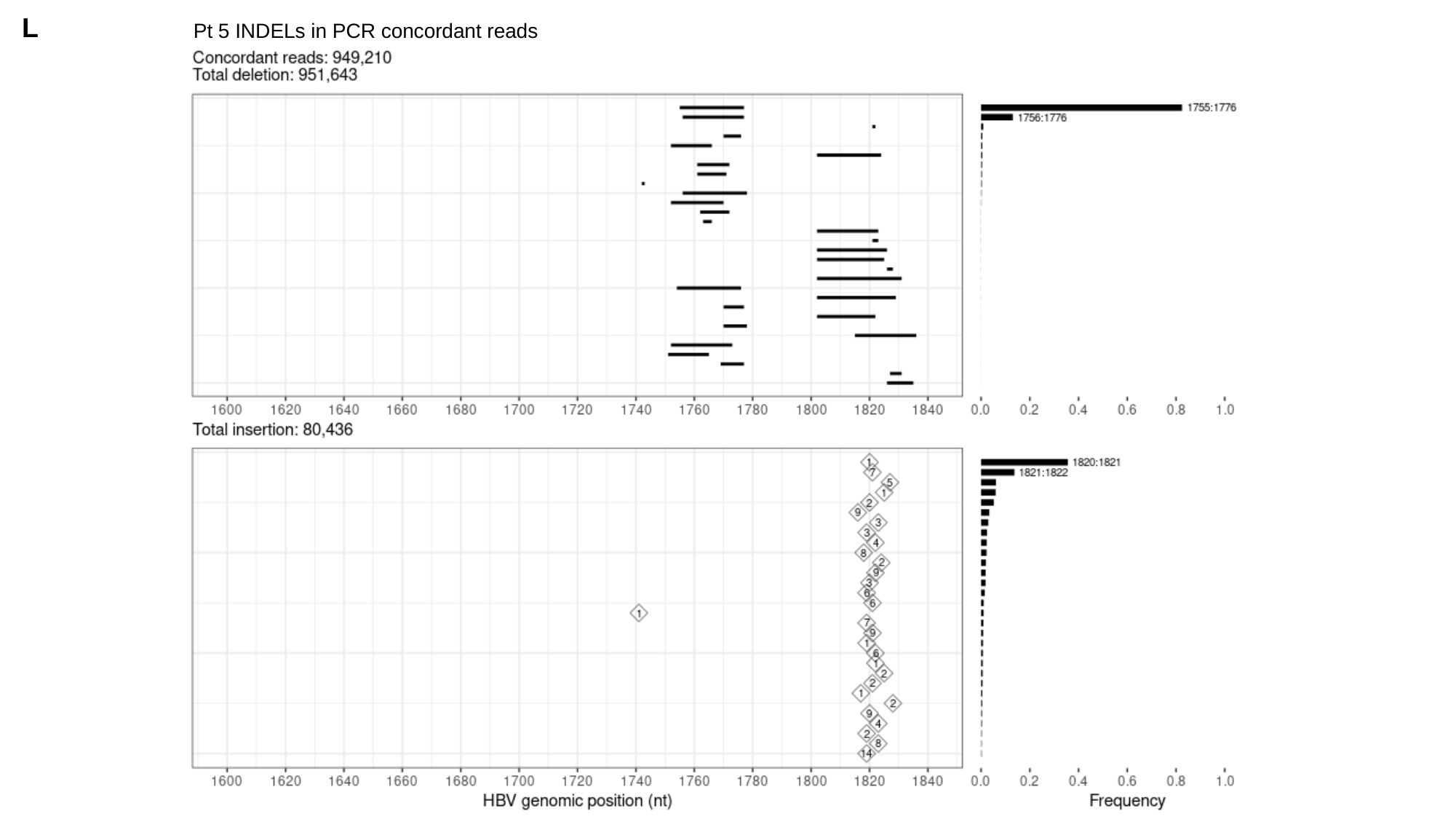

L
Pt 5 INDELs in PCR concordant reads

### Slide 14
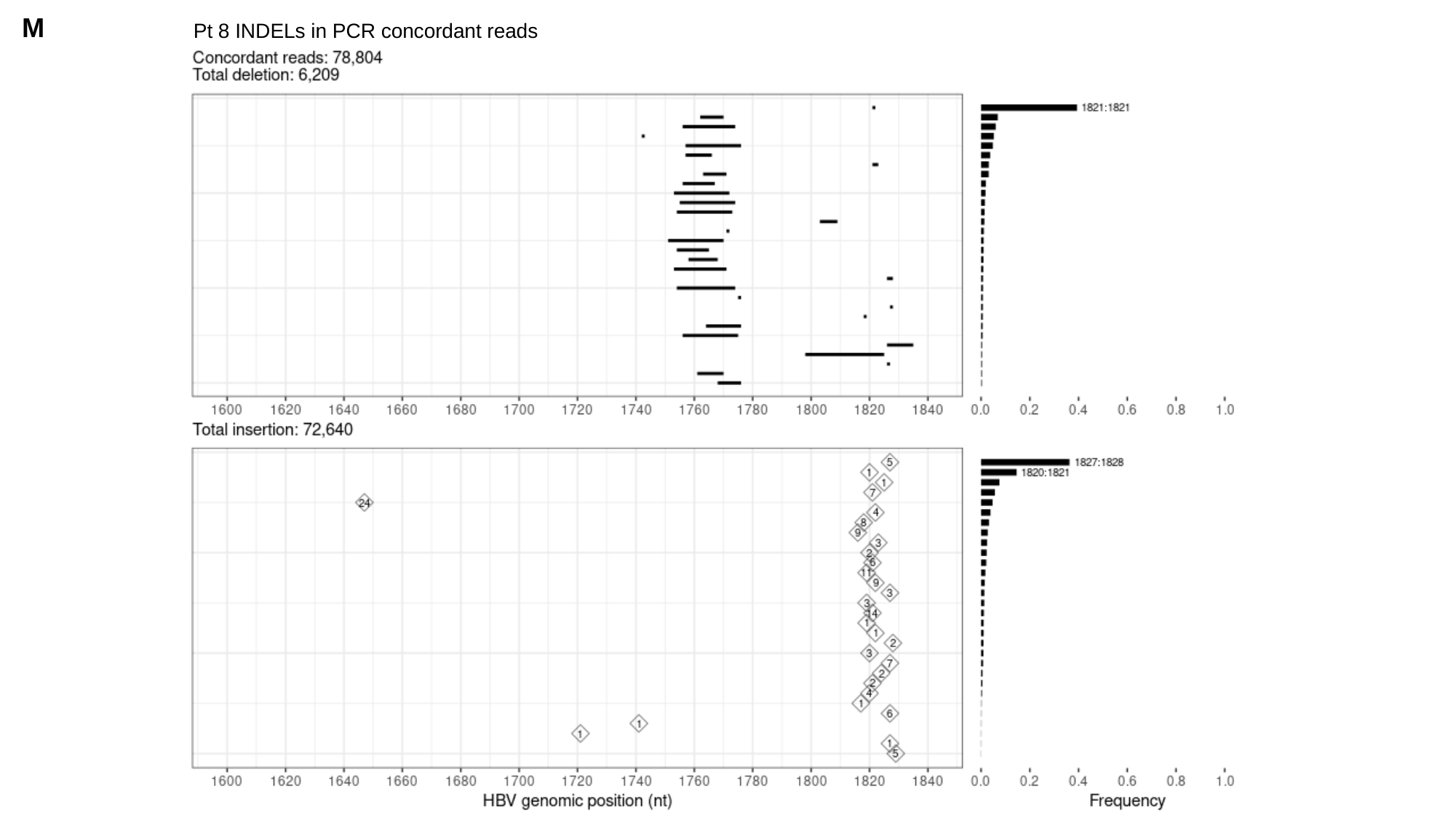

M
Pt 8 INDELs in PCR concordant reads

### Slide 15
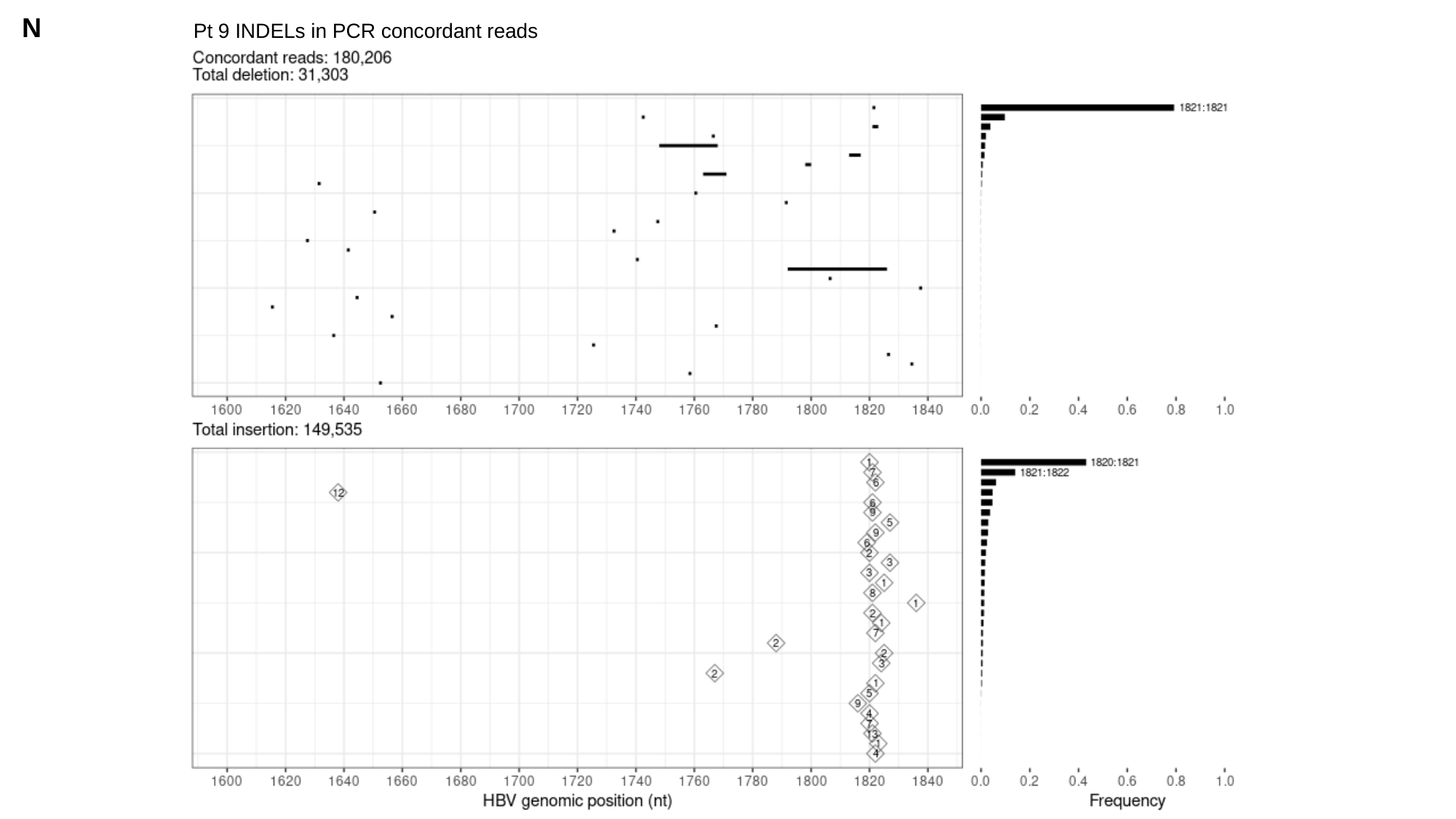

N
Pt 9 INDELs in PCR concordant reads

### Slide 16
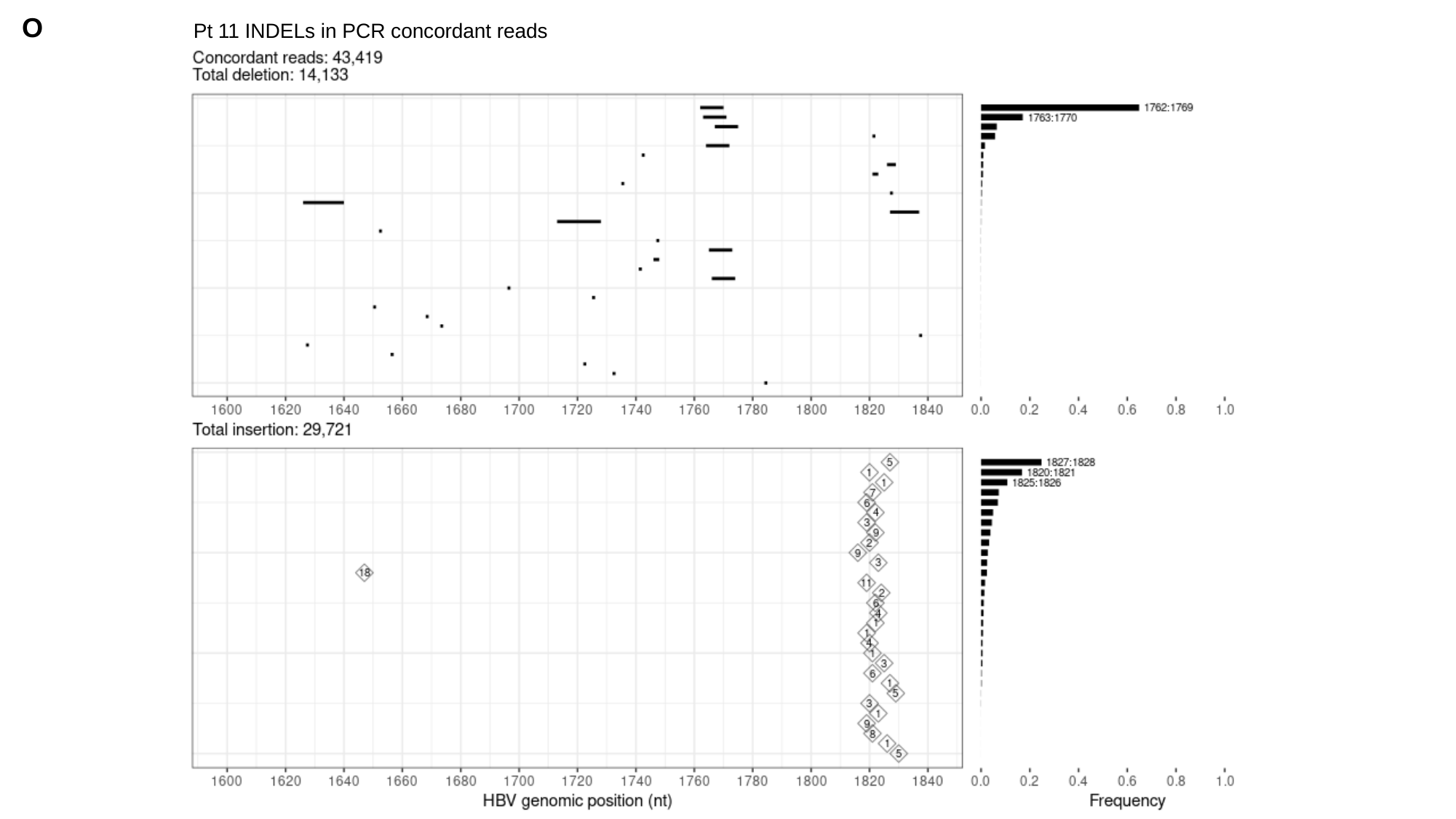

O
Pt 11 INDELs in PCR concordant reads

### Slide 17
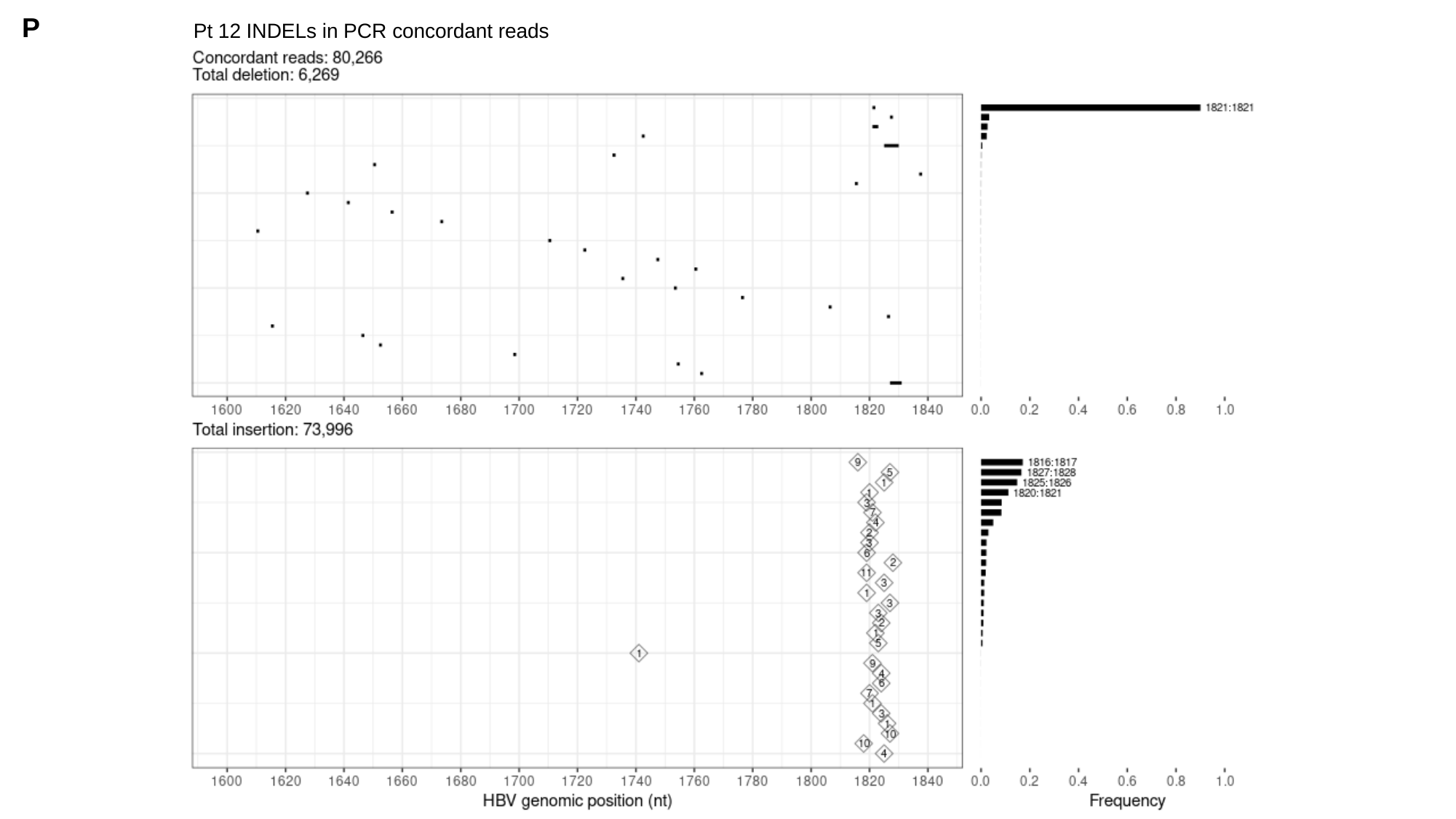

P
Pt 12 INDELs in PCR concordant reads

### Slide 18
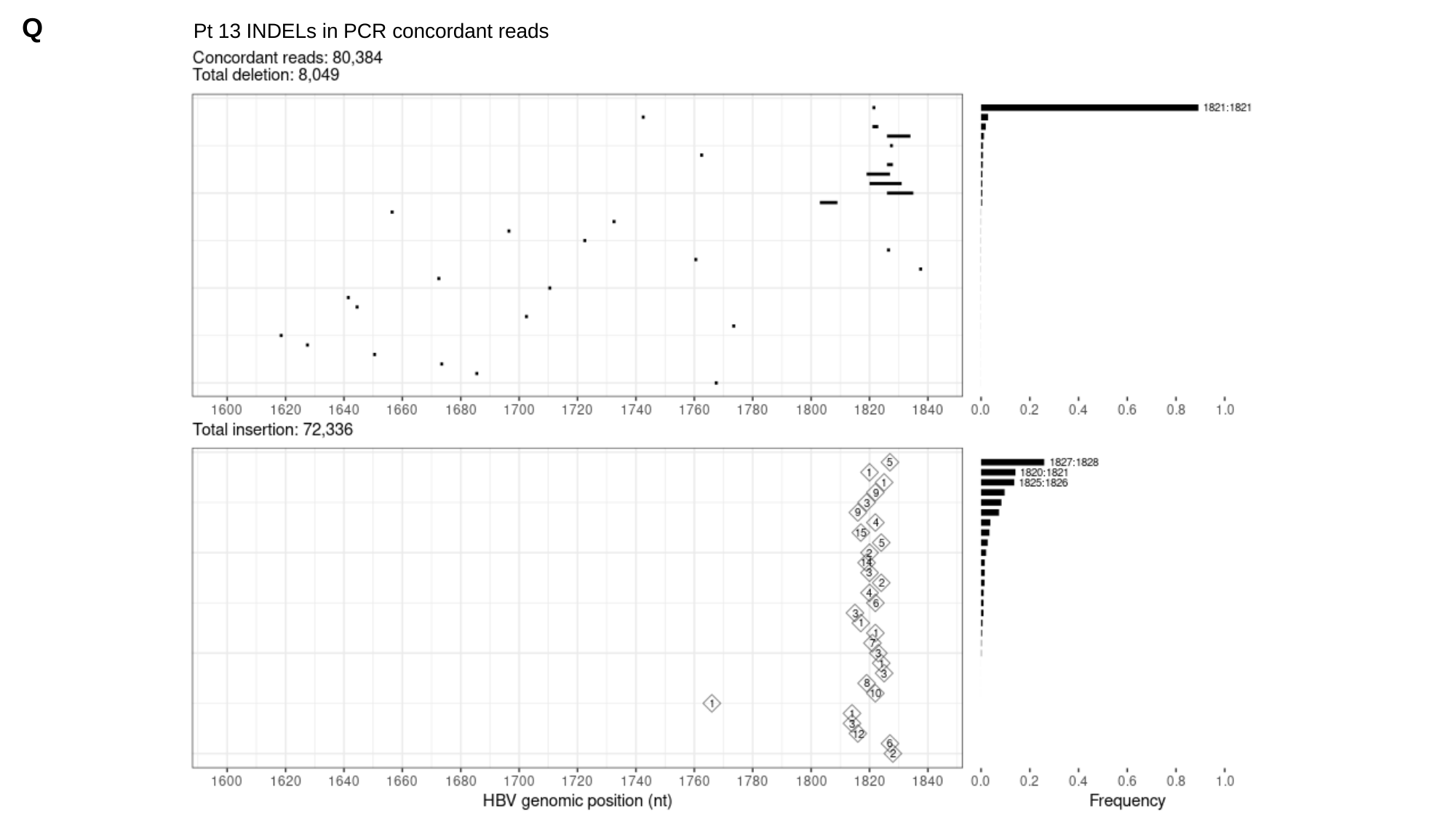

Q
Pt 13 INDELs in PCR concordant reads

### Slide 19
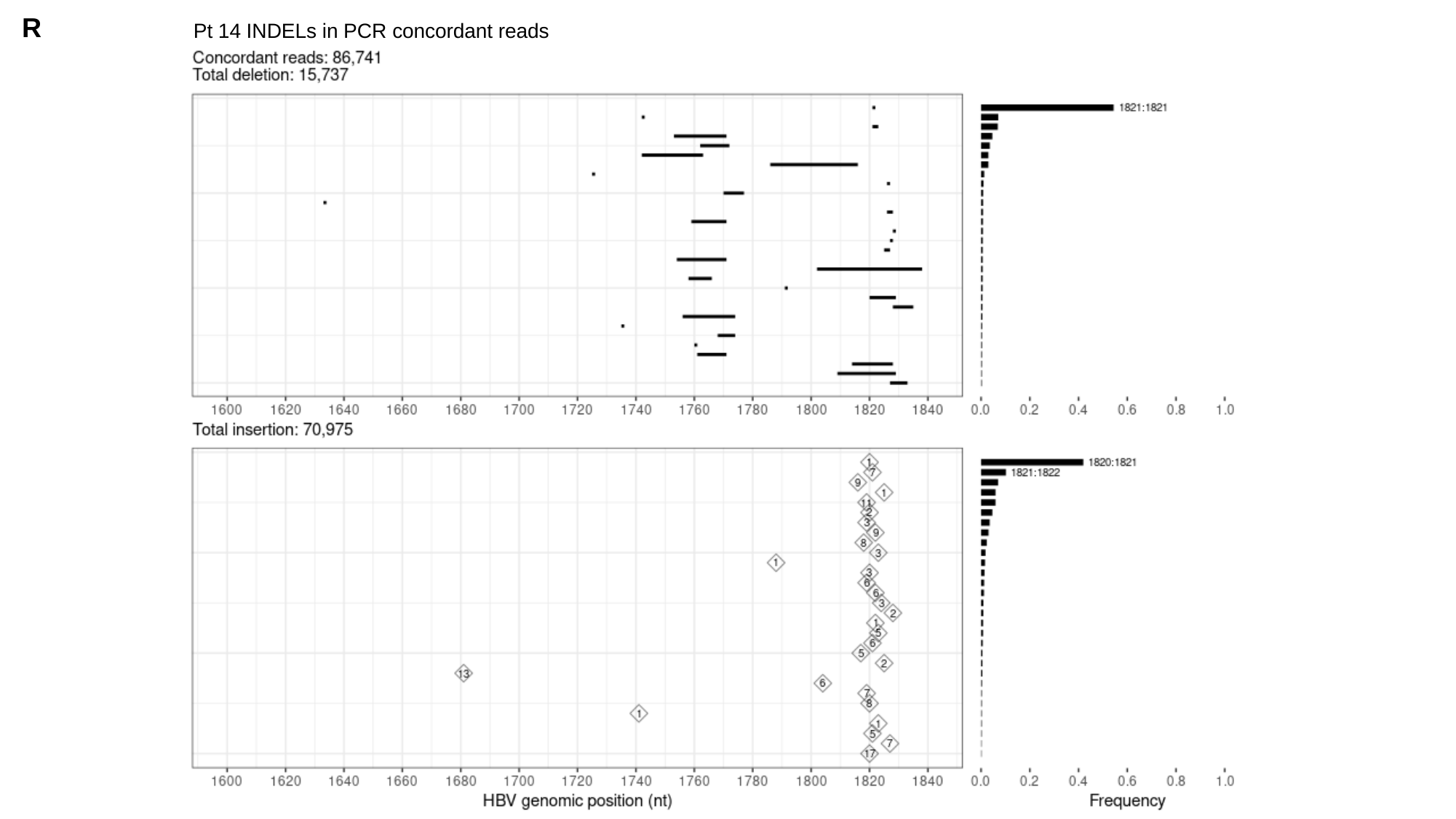

R
Pt 14 INDELs in PCR concordant reads

### Slide 20
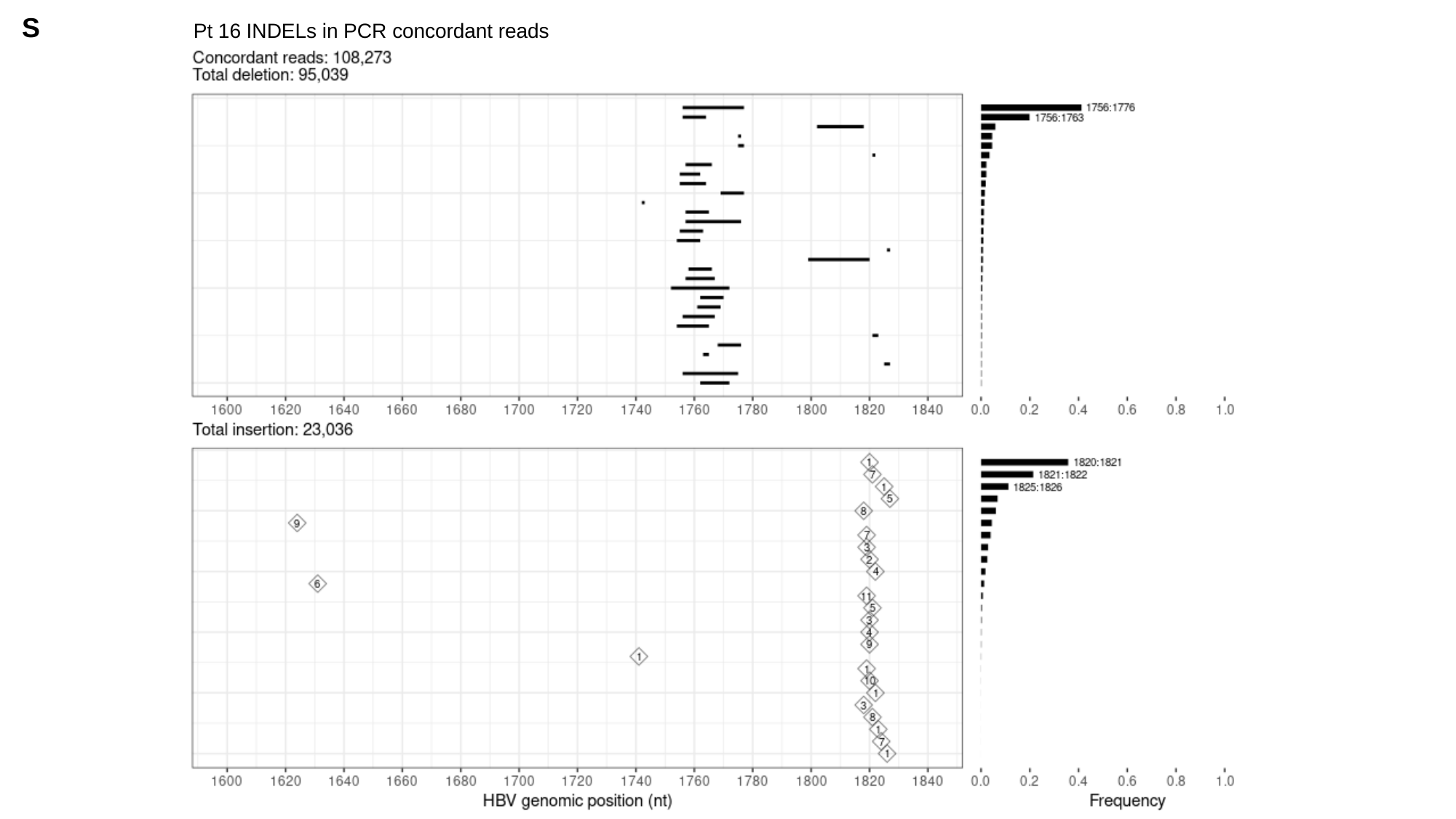

S
Pt 16 INDELs in PCR concordant reads

### Slide 21
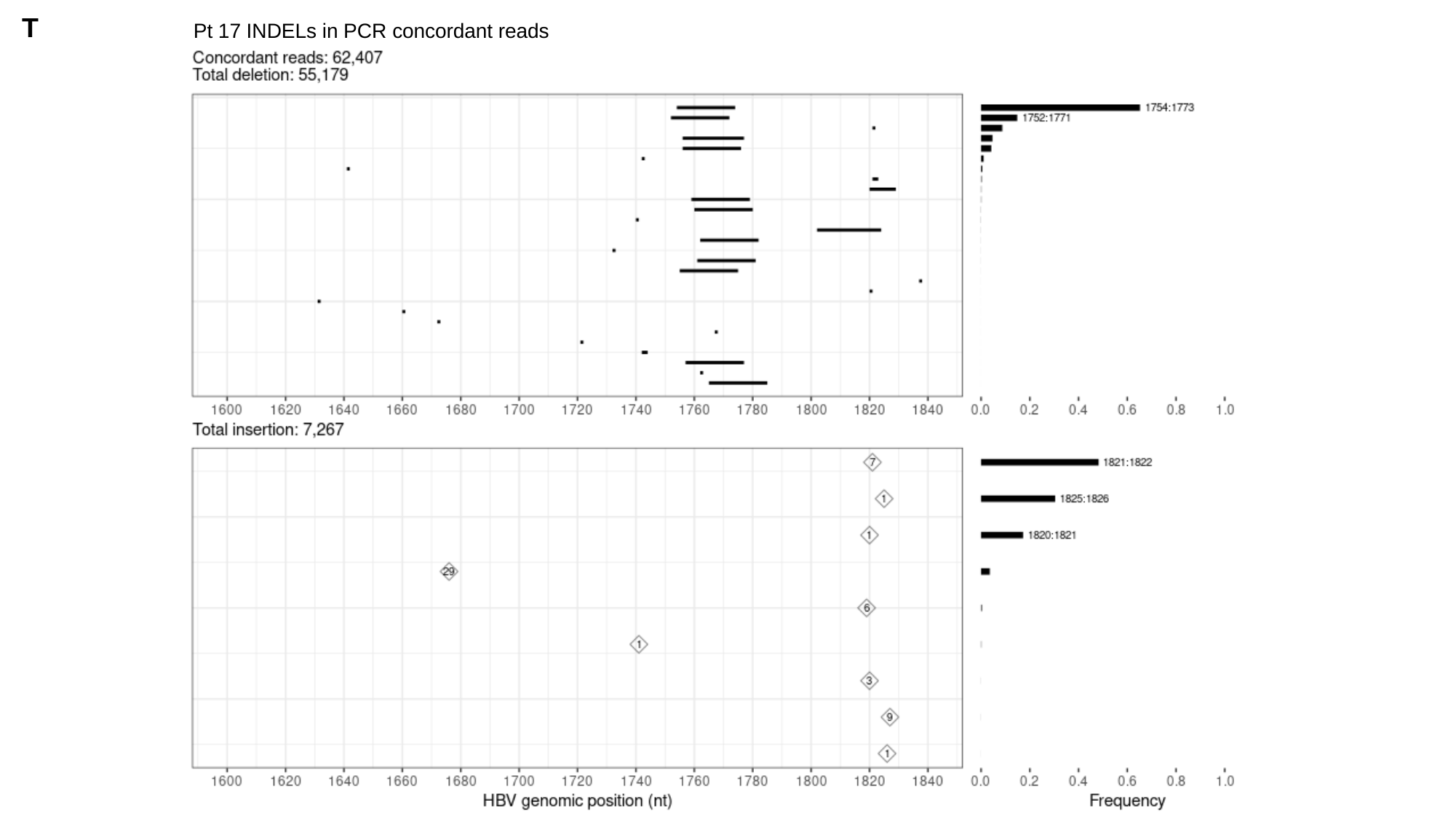

T
Pt 17 INDELs in PCR concordant reads

### Slide 22
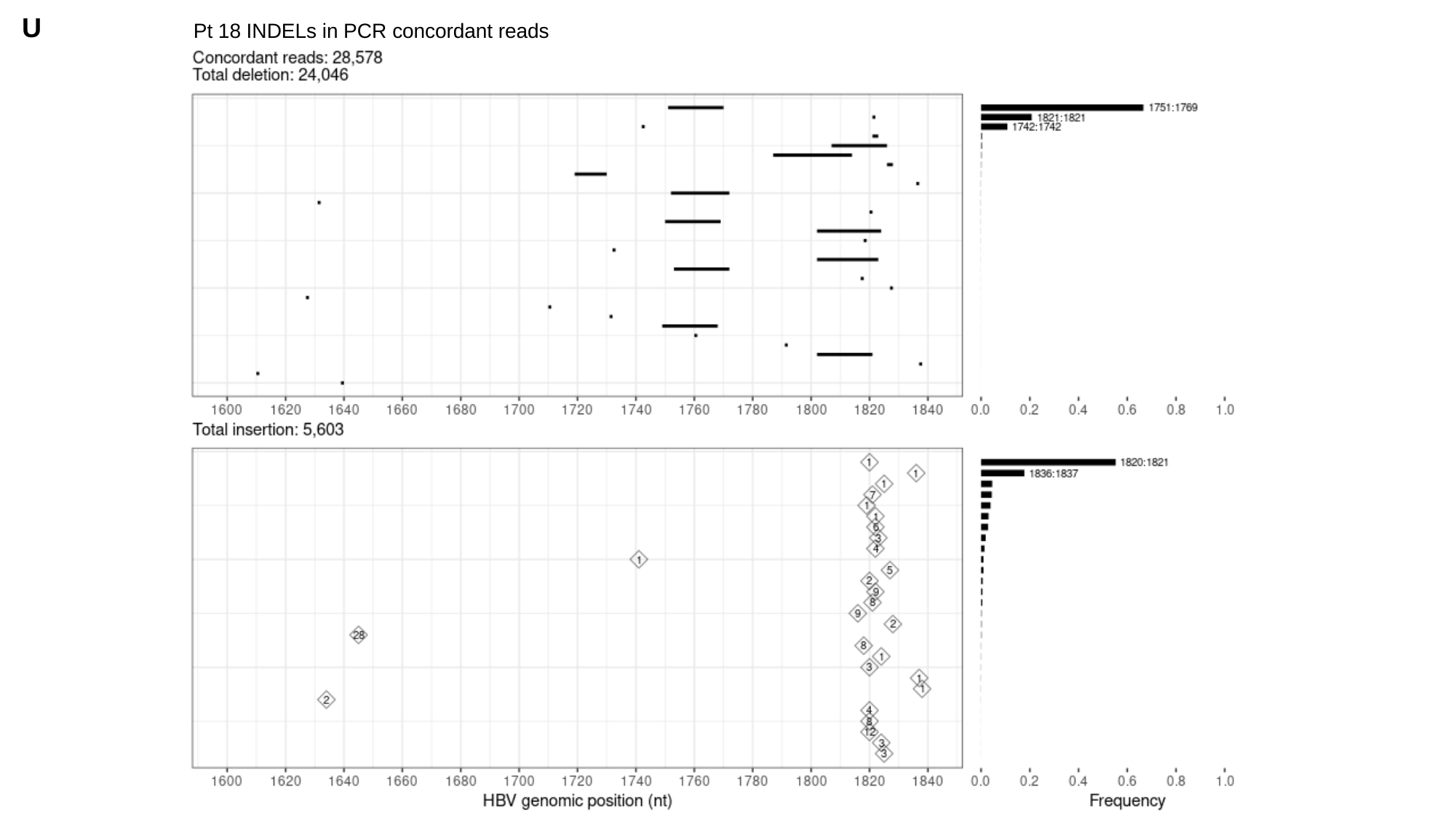

U
Pt 18 INDELs in PCR concordant reads

### Slide 23
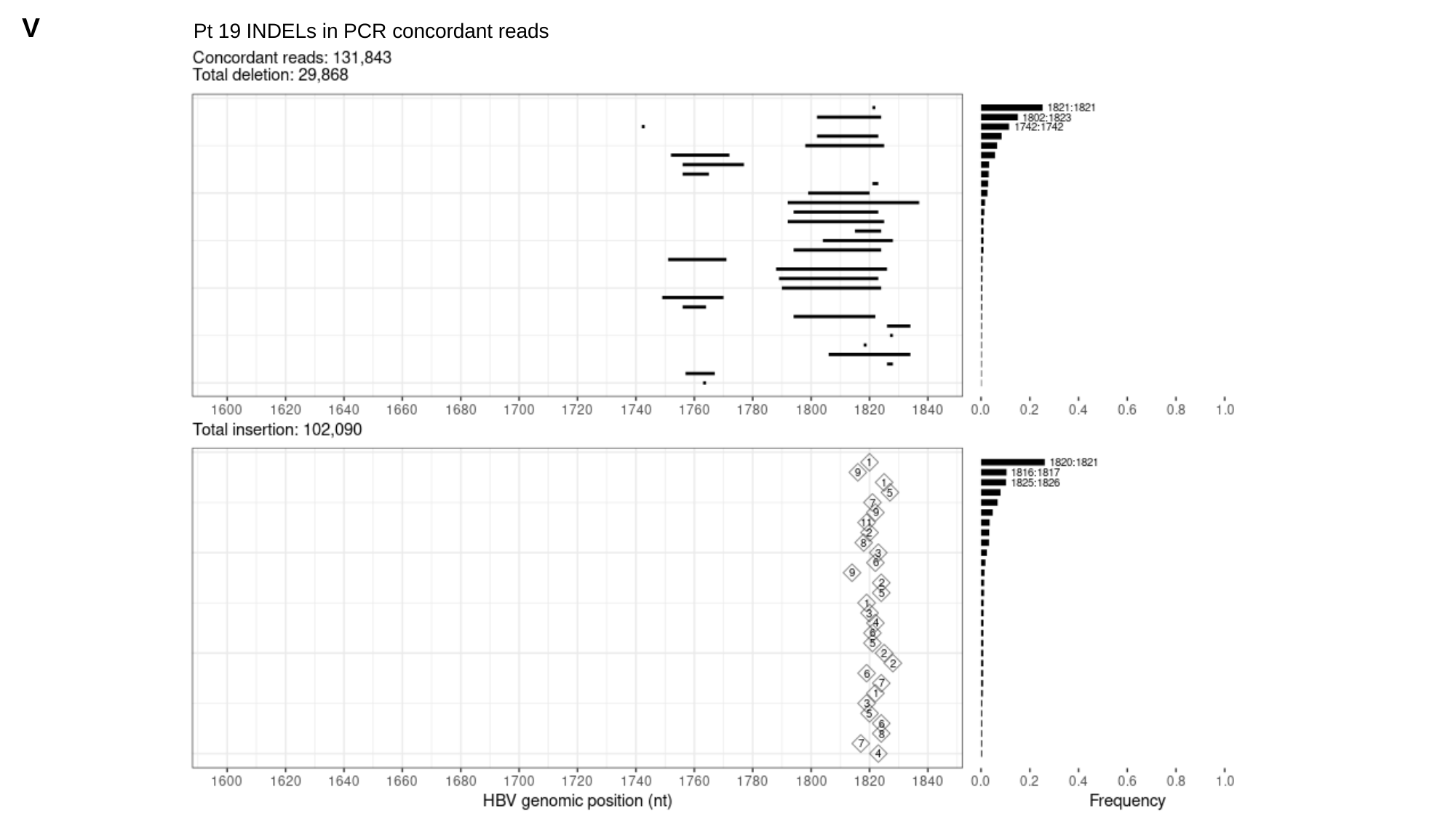

V
Pt 19 INDELs in PCR concordant reads

### Slide 24
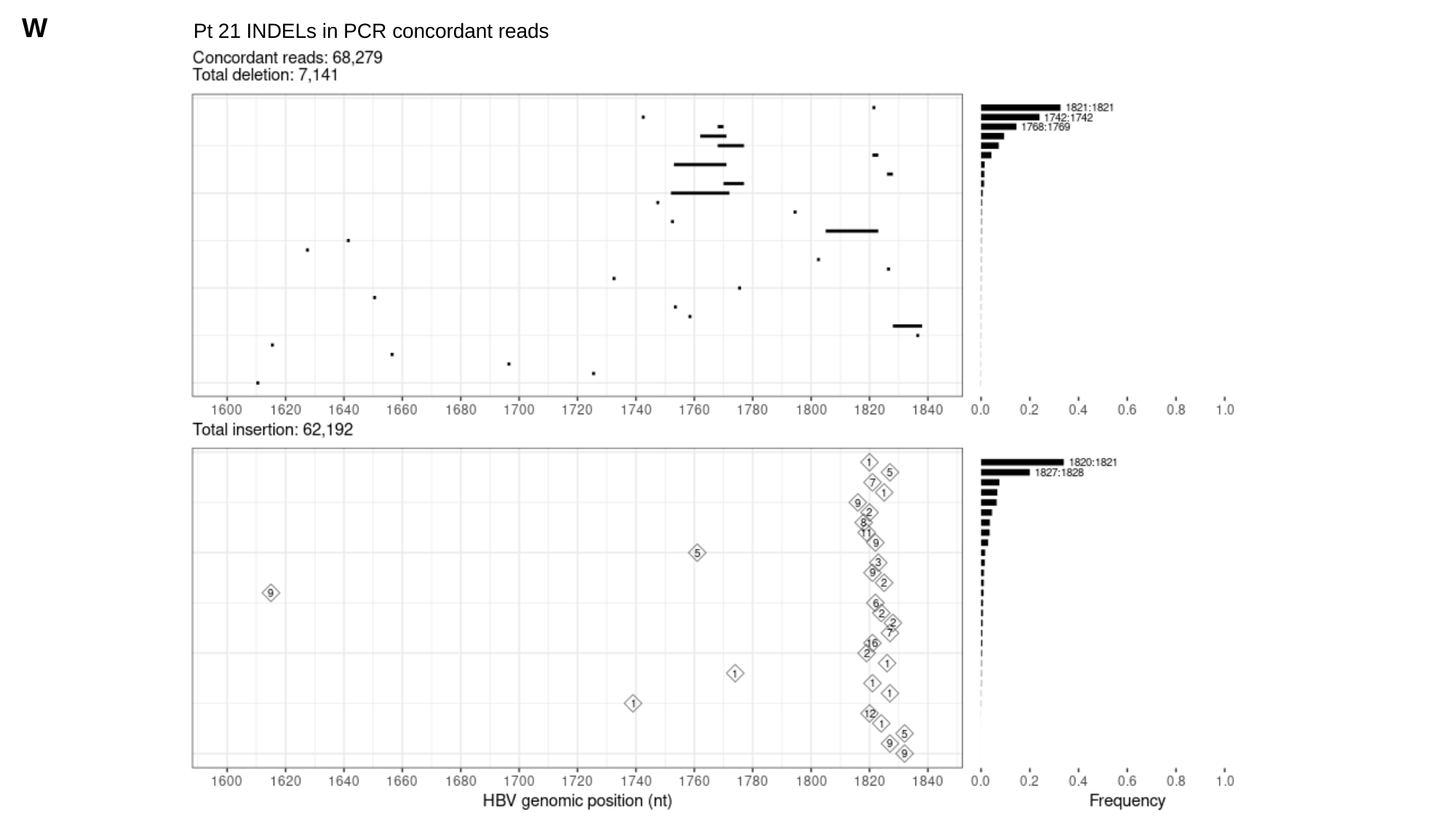

W
Pt 21 INDELs in PCR concordant reads

### Slide 25
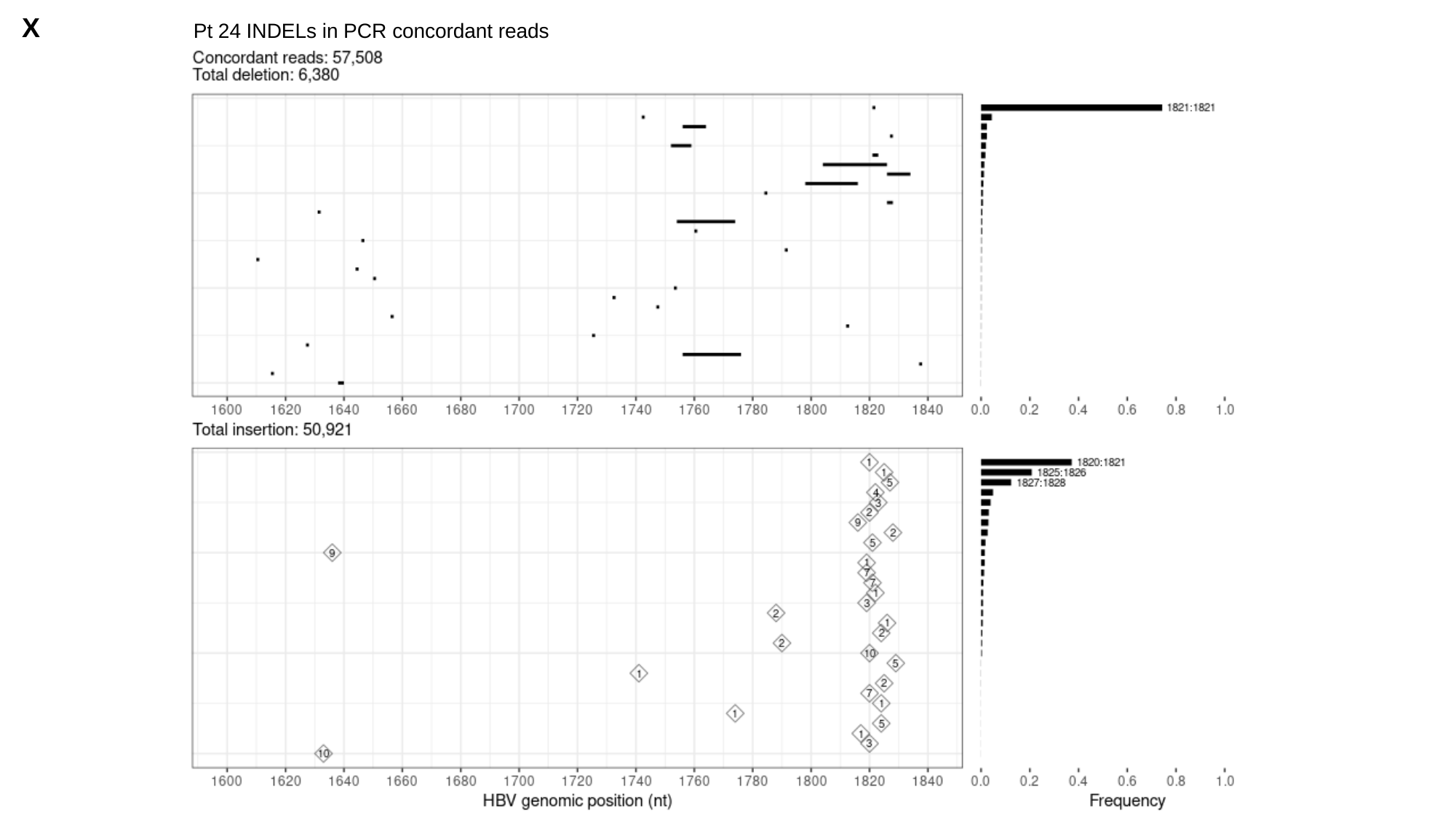

X
Pt 24 INDELs in PCR concordant reads

### Slide 26
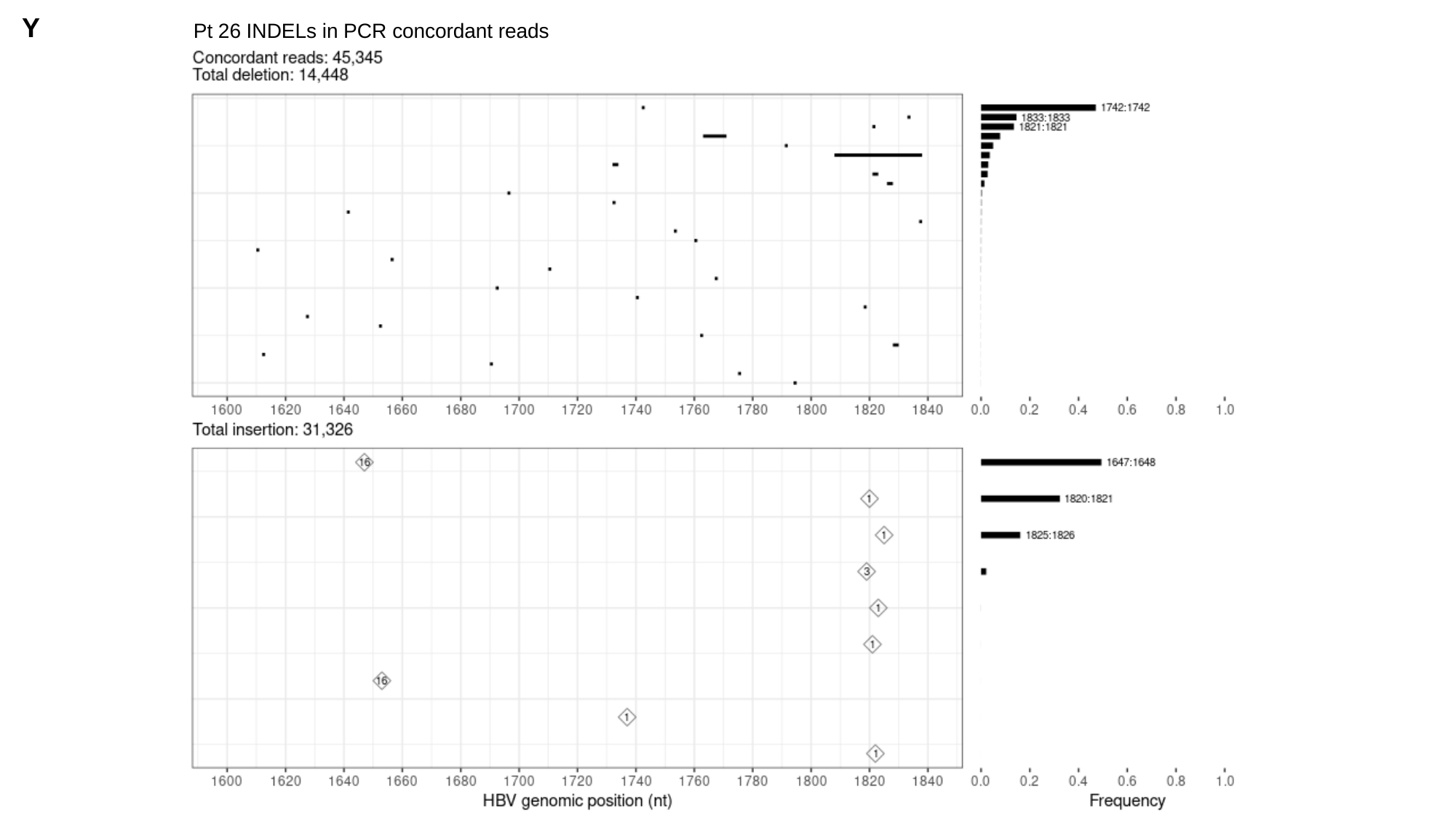

Y
Pt 26 INDELs in PCR concordant reads

### Slide 27
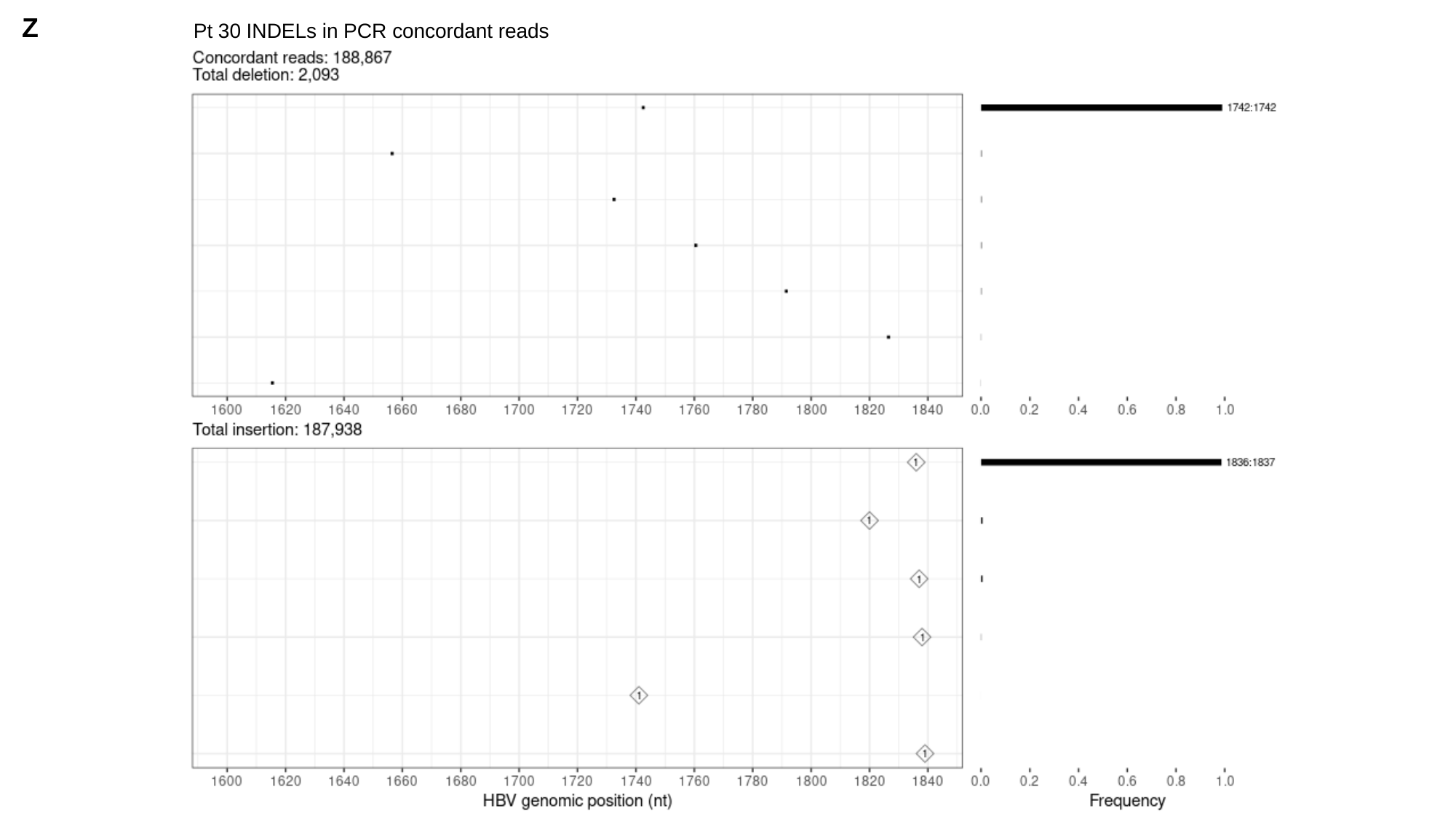

Z
Pt 30 INDELs in PCR concordant reads

### Slide 28
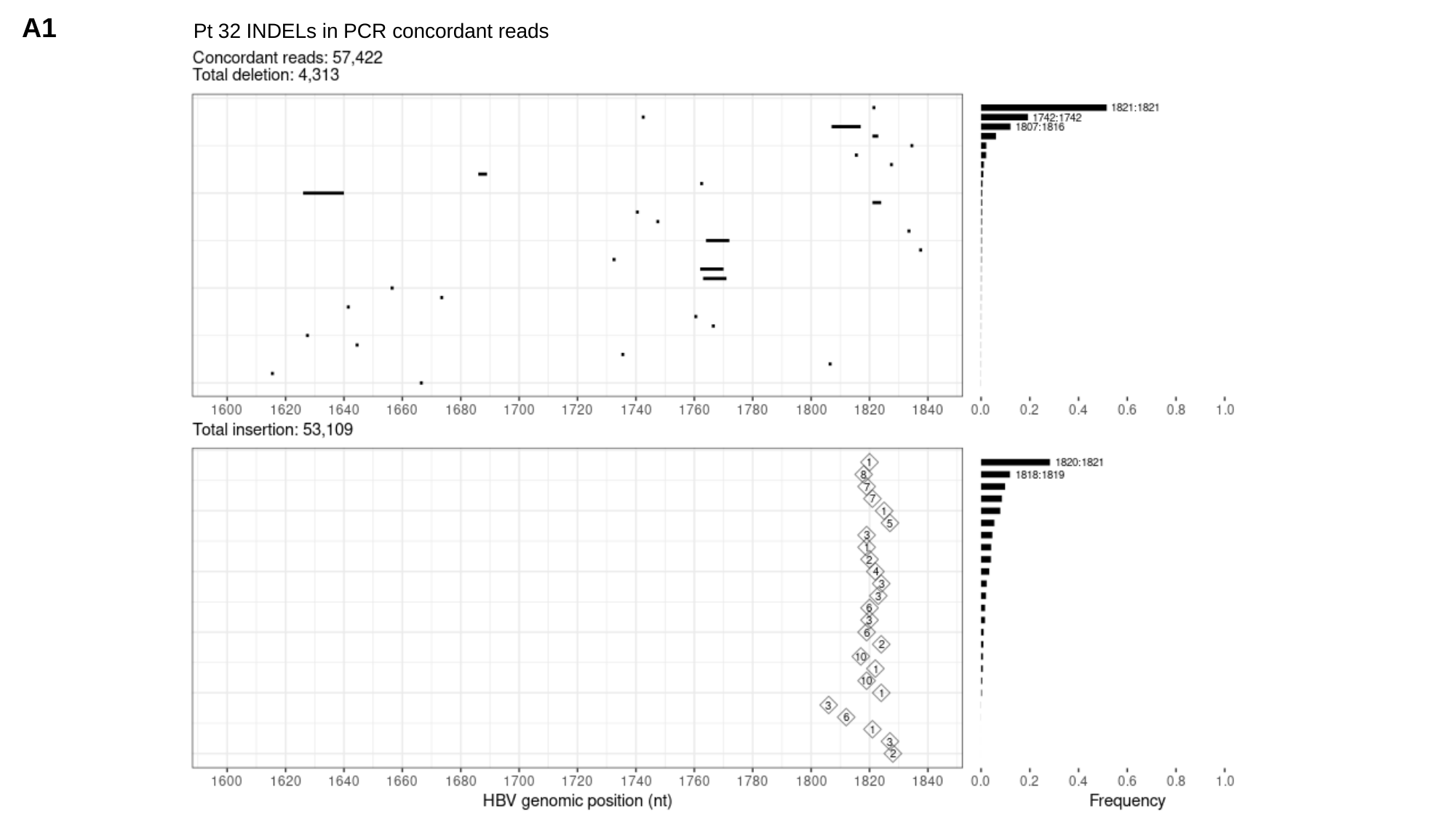

A1
Pt 32 INDELs in PCR concordant reads

### Slide 29
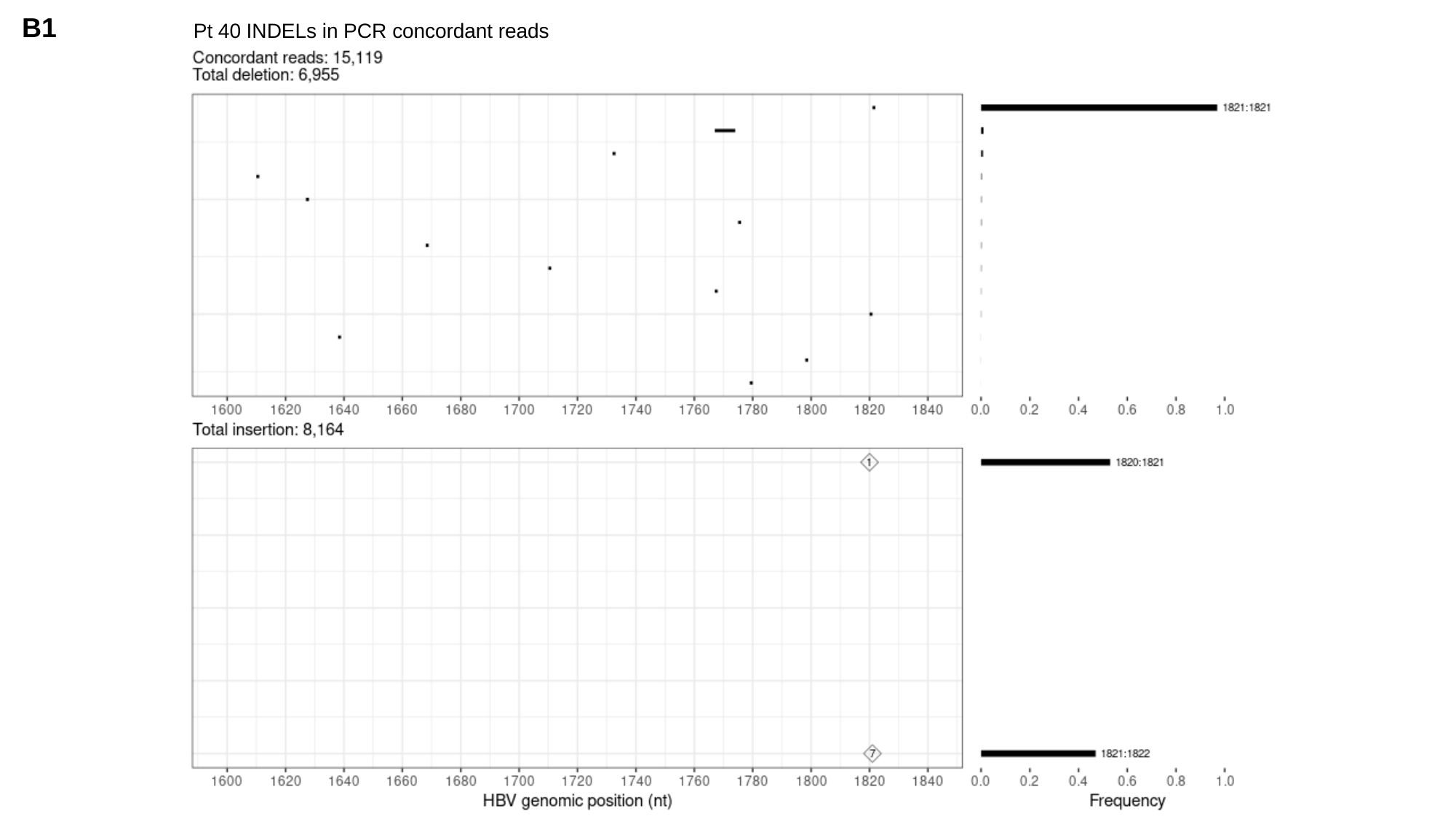

B1
Pt 40 INDELs in PCR concordant reads

### Slide 30
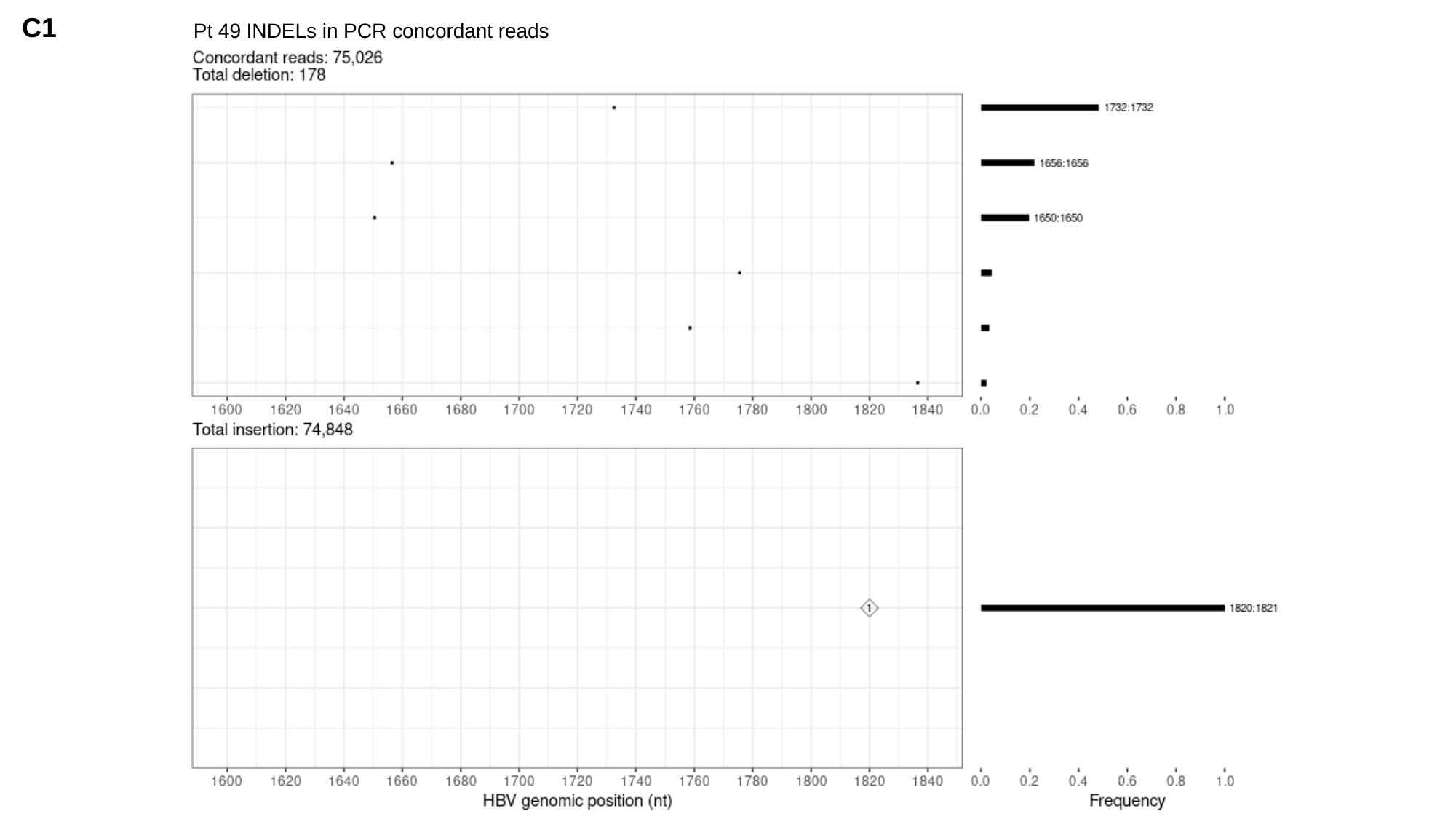

C1
Pt 49 INDELs in PCR concordant reads

### Slide 31
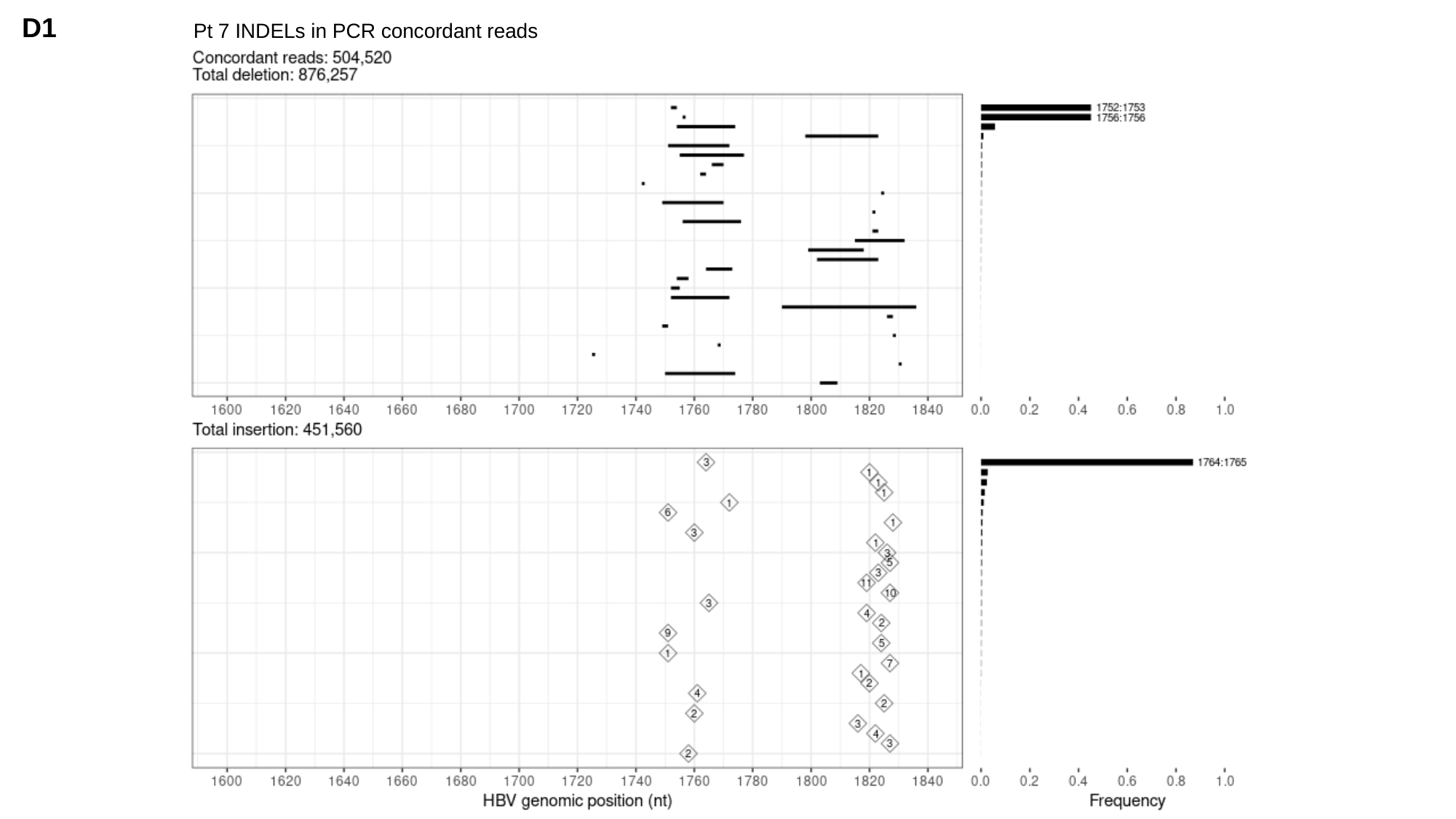

D1
Pt 7 INDELs in PCR concordant reads
